# Drinking water chlorination impact on fecal carriage of extended-spectrum beta-lactamase-producing *Enterobacteriaceae* in Bangladeshi children in a double-blind, cluster-randomized controlled trial

**DOI:** 10.1101/2022.04.04.22273386

**Authors:** Maria Camila Montealegre, Esther E. Greenwood, Lisa Teichmann, Maya L. Nadimpalli, Lea Caduff, Jenna M. Swarthout, Tabea Nydegger, Sonia Sultana, Mohammad Aminul Islam, Val F. Lanza, Stephen P. Luby, Amy J. Pickering, Timothy R. Julian

## Abstract

**Background:** Water, sanitation, and hygiene (WASH) services have the potential to interrupt transmission of antimicrobial-resistant bacteria and reduce the need for antibiotics, thereby reducing selection for resistance. However, evidence of WASH impacts on antimicrobial resistance (AMR) is lacking.

**Methods:** We evaluated extended-spectrum beta-lactamase (ESBL)-producing *Escherichia coli* and ESBL-KESC (*Klebsiella* spp*., Enterobacter* spp*., Serratia* spp., and *Citrobacter* spp.) carriage in the feces of 479 Bangladeshi children under 5 years of age enrolled in a double-blind, cluster-randomized controlled trial of in-line drinking water chlorination in two low-income urban communities in Bangladesh. We additionally assessed the intervention’s impact on circulating beta-lactamase genes in fecal metagenomes and in genomes of fecal ESBL-*E. coli* isolates.

**Findings:** We detected ESBL-*E. coli* in 65% (n = 309) and ESBL-KESC in 12% (n = 56) of enrolled children. We observed no effect of the intervention on the prevalence of ESBL-*E. coli* (relative risk [95% confidence interval] = 0.98 [0.78, 1.23]) when controlling for study site and age. Although ESBL-KESC (0.76 [0.44, 1.29]) was lower among children in the intervention group, the relative risk was not significant. Concentrations of ESBL-*E. coli* (log_10_ CFU/g-wet) were on average [95% confidence interval] 0.13 [-0.16, 0.42] higher in the intervention group and ESBL-KESC (log_10_ CFU/g-wet) were 0.10 [-0.22, 0.02], lower in the intervention group, when controlling for study site and age. Furthermore, the distribution of ESBL-*E.coli* sequence types, type of beta-lactamase-encoding genes in ESBL-*E. coli* isolates, and the presence and relative abundance of beta-lactamase-encoding genes in children’s fecal metagenomes did not differ significantly between the intervention and control children.

**Interpretation:** One year of in-line drinking water chlorination in communities did not meaningfully impact the carriage of ESBL-*E. coli* among children in an area of high ESBL-*E. coli* carriage. While ESBL-KESC was at lower prevalence than ESBL-*E. coli*, in the intervention group, limited study power prevented a clear interpretation of treatment effect. Development and evaluation of effective interventions to reduce AMR carriage are needed to support calls for WASH embedded in current National and Global AMR Action Plans.

## INTRODUCTION

Extended-spectrum beta-lactamase-producing *Enterobacteriaceae* (ESBL-E) impose a burden on human health in hospitals and community settings as infections caused by ESBL-E increase morbidity, mortality, and health treatment costs.^1,2^ Intestinal ESBL-E carriage among healthy individuals, which frequently precede ESBL-E infections,^3^ has shown increasing trends with an estimated annual global growth rate of 5%.^4,5^ The prevalence of ESBL-E carriers differs substantially among regions, with the highest rates reported for South Asia, where 74% of healthy Bangladeshi infants under 1 year of age were reported as carriers of ESBL-*E. coli*.^4,6^ Because ESBL-E are pervasive in humans, animals, and environmental compartments,^7–9^health risks arise from transmission at the interface of these three domains as conceptualized by One Health.^10–13^

Water, sanitation, and hygiene (WASH) services are described in national and international action plans as necessary interventions to curb antimicrobial resistance (AMR), despite a lack of supporting data.^14–16^ For example, improving drinking water quality is one of the pillars in the World Health Organization’s Global Action Plan on AMR to limit the dissemination of antimicrobial-resistant bacteria and reduce human risks associated with environmental exposures.^17^ WASH services are thought to reduce exposure to fecal bacteria, including enteric pathogens, and so may impact AMR by both reducing exposures to antimicrobial-resistant bacteria as well as the use of antibiotics to treat enteric diseases.^15,18^ Although the conceptual model is sound, there is little empirical evidence that WASH services reduce AMR. Indeed, Berendes et al. (2019) found no association between carriage of antimicrobial resistance genes and WASH infrastructure amongst Mozambican children.^19^ Furthermore, several randomized controlled trials of household-level WASH interventions have shown mixed efficacy in reducing environmental fecal contamination, pediatric diarrheal disease, and pediatric growth-faltering and stunting.^20–24^ For WASH interventions to be effective in reducing AMR, the interventions likely need to be effective at reducing transmission of antimicrobial-resistant bacteria and enteropathogens. Given this, automated in-line drinking water chlorination at the point of collection is a promising WASH intervention with potential to reduce AMR. A recent cluster-randomized controlled trial in Bangladesh showed that in-line water chlorination reduced pediatric diarrheal disease by 23%, antibiotic use by 7%, and illness-related expenditures by an average of approximately US$0.50.^25^ Further, *E. coli* prevalence in the tap was reduced (15% in treatment compared to 64% in control) and *E. coli* concentrations were 0.86 log_10_ colony forming units lower in the treatment compared to the control.^25^ The trial results highlight the potential of WASH investments to reduce antibiotic consumption and impact fecal-oral transmission pathways directly relevant to enteropathogen and AMR transmission.

In this study, we assessed ESBL-*E. coli* fecal carriage among children enrolled in the in-line drinking water chlorination cluster-randomized controlled trial. Our primary goal was to evaluate the impact of drinking water chlorination on prevalence and concentration of ESBL-*E. coli* in children’s fecal samples. Secondary analyses included investigating the intervention’s impact on carriage of ESBL-KESC (*Klebsiella* spp*., Enterobacter* spp*., Serratia* spp., and *Citrobacter* spp.), the presence/absence and relative abundance of beta-lactamase genes in children’s fecal metagenomes and in genomes of fecal ESBL-*E. coli* isolates.

## MATERIALS AND METHODS

### Study design and fecal samples

We analyzed 479 fecal samples of children under 5 years of age previously enrolled in a double-blind, cluster-randomized controlled trial of in-line water chlorination conducted in two low-income communities in Bangladesh (Dhaka and Tongi) between July 2015 and December 2016 (Table 1).^25^ Our study was powered to detect a 12% reduction in ESBL-*E. coli* carriage rates with 80% power at 5% significance, based on a planned sample size of 600 samples collected from 80 clusters (40 intervention, 40 control), cluster coefficient of variation of 0.2, and carriage rate of ESBL-*E. coli* of 30% in the control group.^26,27^ Clusters were defined as all households with a child under five that access a single water point, and contained one or more compounds, defined as a set of households owned by a single landlord with shared facilities. The sample size (n = 479) was lower than the planned sample size (n = 600) because the parent trial ended earlier than desired due to a shortage of chlorine refills (Pickering et al.) and not all planned stool samples were collected. Among the samples collected, an approximately equal number of samples were analyzed from the treatment group as compared to the control.

**Table 1.** Demographics and study characteristics of control (n = 239) and intervention (n = 240) groups, IQR is the interquartile range for length of child enrollment.

|  |  | Control | Intervention |
| --- | --- | --- | --- |
| Study site in Dhaka | *percent* | 33% | 48% |
| Water tap in compound or home | *percent* | 95% | 98% |
| Regularly treats drinking water | *percent* | 16% | 18% |
| Single household compound | *percent* | 19% | 20% |
| People in household | *mean (±SD)* | 4.7 (±1.8) | 4.7 (±1.7) |
| Households in compound | *mean (±SD)* | 8.7 (±10) | 14 (±17) |
| Length of child enrollment | *median [IQR] days* | 330 [267, 346] | 322 [235, 338] |
| Children enrolled < 180 days | *percent* | 13.3% | 18.5% |

The enrolled children used shared water points as their primary drinking water source, at which passive dosing devices were installed and randomly assigned to supply chlorine (intervention group) or vitamin C (active control group).^25^ Vitamin C was dosed in the active control to provide a benefit to study participants, increasing study acceptability, and because it was available in tablets compatible with the chlorine dosing device.^25^ Supplementation with Vitamin C has no documented impact on antimicrobial resistance carriage, though doses substantially larger than those provided here have been shown to impact gut microbiota of animals and humans in small sample trials. ^28–30^ Due to high urban migration, the trial followed an open cohort design, and therefore the time each child was enrolled in the study and exposed to the intervention varied.^25^ Study enrollment was a mean of 9.3 (median 10.7) months. Notably, 73 children (15.9%) were enrolled for fewer than 6 months. The raw fecal samples were stored at −80°C for up to three years at icddr,b (Dhaka, Bangladesh) and shipped on dry ice to Eawag (Dübendorf, Switzerland) for processing. The study was conducted double-blind: neither the study participants nor researchers processing the samples and assessing the study outcomes were aware of which children were receiving chlorinated water.

### Ethical Considerations

The study protocol for the original trial^25^ was approved by the International Centre for Diarrhoeal Diseases Research, Bangladesh (icddr,b) scientific and ethical review committees (protocol number 14022) and the human subjects institutional review board at Stanford University (protocol number 30456). Within the original trial, informed written consent was obtained from all study participants as well as the owners of the water points enrolled. Consent included biospecimen collection for future unplanned analyses.^25^ This study and the original trial follow the Consort 2010 checklist (Supplementary Information).

### Enumeration of E. coli and ESBL-E from feces

We enumerated ESBL-*E. coli* and ESBL-KESC directly from fecal samples using CHROMID^®^ ESBL agar (bioMérieux, Marcy-L’Étoile, France). In brief, we suspended 0.1 ± 0.04 g of each stool sample in 0.9 ml of 0.9% NaCl-solution (10^-1^ dilution of the sample), vortexed for 1 min at maximum speed, and centrifuged at 100 x *g* for 30 s. We used 0.1 ml of the resulting supernatant to prepare 10-fold serial dilutions (10^-2^ −10^-4^) in 0.9% NaCl-solution. We spread-plated the dilutions onto the selective ESBL agar, incubated overnight at 37°C, and enumerated using the reference color indicated by the manufacturer, based on the detection of ß-glucuronidase for ESBL-*E. coli* and ß-glucosidase for ESBL-KESC.^31^ We calculated the colony-forming units per gram of wet feces (CFU/g) from plated dilutions with up to 250 CFU, prioritizing plates with 25-250 CFU.^32^ From a subset of samples, we also directly enumerated ß-glucuronidase *E. coli* from CHROMagar^TM^ orientation (CHROMagar, Paris, France) (n= 113). The upper limit of quantification (ULOQ) was assigned to samples with more than 250 *E. coli* or KESC CFU on the 10^-4^ dilution, corresponding to 6.3 log_10_ CFU/g-wet feces.

We further inoculated 0.1 ml of the supernatant from the centrifuged fecal suspension into 0.9 ml of non-selective tryptic soy broth to enrich the samples and improve detection of ESBL-E. The inoculated trypic soy broth was incubated overnight at 37°C. For samples with detectable ESBL-E colonies on the CHROMID^®^ ESBL agar plates from the dilutions, the enrichment broths were discarded. For samples without detectable ESBL-E colonies, 0.1 ml of the enrichment broth was spread-plated onto selective CHROMID^®^ ESBL agar plates. When the plated enrichments were positive for *E. coli* or KESC, the lower limit of detection (2 log_10_ CFU/g-wet feces) was assigned to the sample. The lower limit of detection was determined by multiplying the lowest detectable number of colony forming units per plate (1) by the lowest dilution plated for the sample (10^-2^) and dividing by the lowest wet fecal weight used across all samples (0.1 g). When both the plated dilutions and plated enrichments were negative for *E. coli* or KESC, the sample was considered negative and a value of half the lower limit of detection (1.7 log_10_ CFU/g-wet feces) was assigned to the sample for quantitative analysis. Due to differences in weighted grams of stool per sample, some calculated values were outside of the range of the lower limit of detection or upper limit of quantification values. These values were replaced with the corresponding lower limit of detection or upper limit of quantification.

### Statistical analyses of the intervention’s impact on ESBL-E. coli and ESBL-KESC

The impact of the intervention on the detection of culturable ESBL-*E. coli* (primary outcome) and ESBL-KESC were assessed using modified Poisson regression, controlling for study area categorized by the two communities included in the study (Dhaka, Tongi) and child age group (<16 months, 16-30 months, >30 months). Age range categories approximately correspond to distinct gut microbiome phases: developmental, transitional, and stable.^33^ The impact of the intervention in the log_10_ CFU/g-wet feces concentrations of ESBL-*E. coli* and ESBL-KESC was assessed using linear regression models, also controlling for the study area and age group. Impact of additional risk factors (gender, study area, children’s age, antibiotic use in the last two months, visit to a treatment facility in the last two months, number of people in the household, and number of households in a compound) on fecal carriage and concentration of ESBL-*E. coli* and ESBL-KESC among children were further explored using Poisson regression (carriage) or linear regression models (concentration). Subgroup analyses for study site and study enrollment duration longer than 10 months were also conducted to investigate effect modification of these risk factors on the intervention impacts. Subgroup analyses for study enrollment duration censored enrollment of children less than 10 months old at the time of sampling, and were therefore performed using binary age categories of less than or equal to 30 months and greater than 30 months old. These and the following statistical analyses were performed using R, version 4.04, R Studio v. 1.1.463, or Prism 8. Statistical significance was defined by α = 0.05.

### Whole-genome sequencing of ESBL-E. coli isolated from feces

We isolated and sequenced the genomes of a total of 96 ESBL-*E. coli* isolates from 86 random child fecal samples, balanced between the control (n = 43) and intervention (n = 43) groups. Ten isolates were from eight random child fecal samples of which one (n = 6) or two (n = 2) additional isolates were sequenced. The bacterial DNA was extracted with the DNeasy Blood & Tissue Kit (Qiagen, Hilden, Germany) and sent to Novogene (UK) Company Limited for sequencing using the Illumina Novaseq 6000 platform (2 x 150 bp) (Illumina, San Diego, USA). The bioinformatic pipeline followed the primary steps as previously described by Montealegre et al..^34^ In brief, Trimmomatic version 0.39 ^35^, SPAdes version 3.11.1^36^, Quast^37^, and Prokka^38^ 1.12 were used to assess read quality, de novo assemble, assess assembly quality, and annotate the genomes, respectively. Multilocus sequence typing (MLST) using the Achtman scheme and phylogenetic group determination were performed in silico using MLST v. 2.16.1 (https://github.com/tseemann/mlst) and Clermontyping (http://clermontyping.iame-research.center). Core genome single nucleotide polymorphisms and indels were identified using Snippy 4.0.^39^ Phylogenetic tree reconstruction by IQ tree (http://www.iqtree.org/), and visualized using ITOL version 4.3.3 (https://itol.embl.de). ABRicate^40^ with the ResFinder^41^ and Virulence Factor^42^ databases were used to identify antimicrobial resistance genes and putative virulence factors (query date 2020-02-10; cutoffs for identity and coverage were >90%). The phylogroup distribution of the isolates (based on the Clermont typing A, B1, B2, C, D, E, F and clade I) was compared between the intervention and the control using Chi-squared tests. ^43^ Similarly, the frequencies of isolates with specific AMR genes or putative virulence factors were compared between the intervention and control using t-tests corrected for family-wise error using the Benjamini-Hochberg method.^44^

### Metagenomics

We characterized the occurrence and relative abundance of beta-lactamase genes in fecal samples using short-read metagenomic sequencing on a subset of 97 fecal samples from children older than 6 months in the control (n=50) and intervention groups (n=47). The subset was chosen from the samples randomly after stratification by intervention group (intervention, control) and study site to ensure balanced sample sizes. Total DNA was extracted from approximately 0.25 g of frozen feces using the QIAamp PowerFecal DNA Kit (Qiagen) according to the manufacturer’s instructions. Total DNA was quantified using a Qubit 2.0 fluorometer (Thermo Fisher Scientific), then sent to Novogene (UK) Company Limited for short-read, paired-end 150 bp sequencing on a Novaseq S4 instrument to achieve 10 Gb per sample. We tabulated the number of raw reads for each sample and excluded from further analysis samples with fewer reads than two standard deviations below the mean (n = 2), resulting in subsequent analysis of 95 fecal samples. We screened for antibiotic resistance genes by mapping raw reads to the Resfinder database (v. 3.1.1) using the KMA tool.^41,45^ We considered matches with >90% coverage and >95% identity to be true hits. We used logistic regression models to examine differences in the presence/absence of beta-lactamase gene alleles between intervention and control children while controlling for age (6-30 months, >30 months) and study site (Dhaka, Tongi). In contrast to prior analyses, the age group 6-30 months combined the developmental (<16 months) and transitional (16-30 months) phase age groups because there were few developmental phase samples sequenced. Multiple comparison correction was performed using Benjamini-Hochberg method.^44^ We also used the *corncob* package in R to identify differences in the relative abundance of beta-lactamase gene alleles, while controlling for age (6-30 months, >30 months) and study site (Dhaka, Tongi)^46^. For both analyses, we only examined differences in genes that occurred in at least 10% of samples.

## RESULTS

The fecal samples analyzed were equally balanced between the intervention (n = 240, 44 clusters) and the active control (n = 239, 43 clusters) groups and the distribution of gender and age did not differ significantly between the groups. More samples were analyzed from the study site in Tongi (n = 283) than in Dhaka (n = 196) (Table 1).

### Fecal carriage of ESBL-E

We detected ß-glucuronidase-producing ESBL-*E. coli* in 65% (n = 309) and ß-glucosidase-producing ESBL-KESC in 12% (n = 56) of the children’s fecal samples (Fig. 1A). Co-carriage of ESBL-*E. coli* and -KESC was observed in 10% (n = 47) of the samples. Amongst samples with quantifiable positive cultures for ESBL-*E. coli* (59% or n = 284) we detected mean concentrations (± standard deviation, SD) of 4.1 (± 1.4); amongst samples with quantifiable positive cultures for ESBL-KESC (11% or n = 52), mean (SD) concentrations were 3.3 (± 1.2) log_10_ CFU/g-wet feces (Fig. 1B). Amongst samples without quantifiable positive cultures for ESBL-*E. coli,* 77% (n = 151) were negative by enrichment, 13% (n = 26) were positive by enrichment, and 9% (n = 18) were not enriched and so assigned as negative. Concentrations of ESBL-*E. coli* exceeded the upper limit of quantification (6.3 log_10_ CFU/g-wet feces) in 6% (n = 30) of all samples and for ESBL-KESC in 0.4% (n = 2) of all samples.

**Figure 1.**
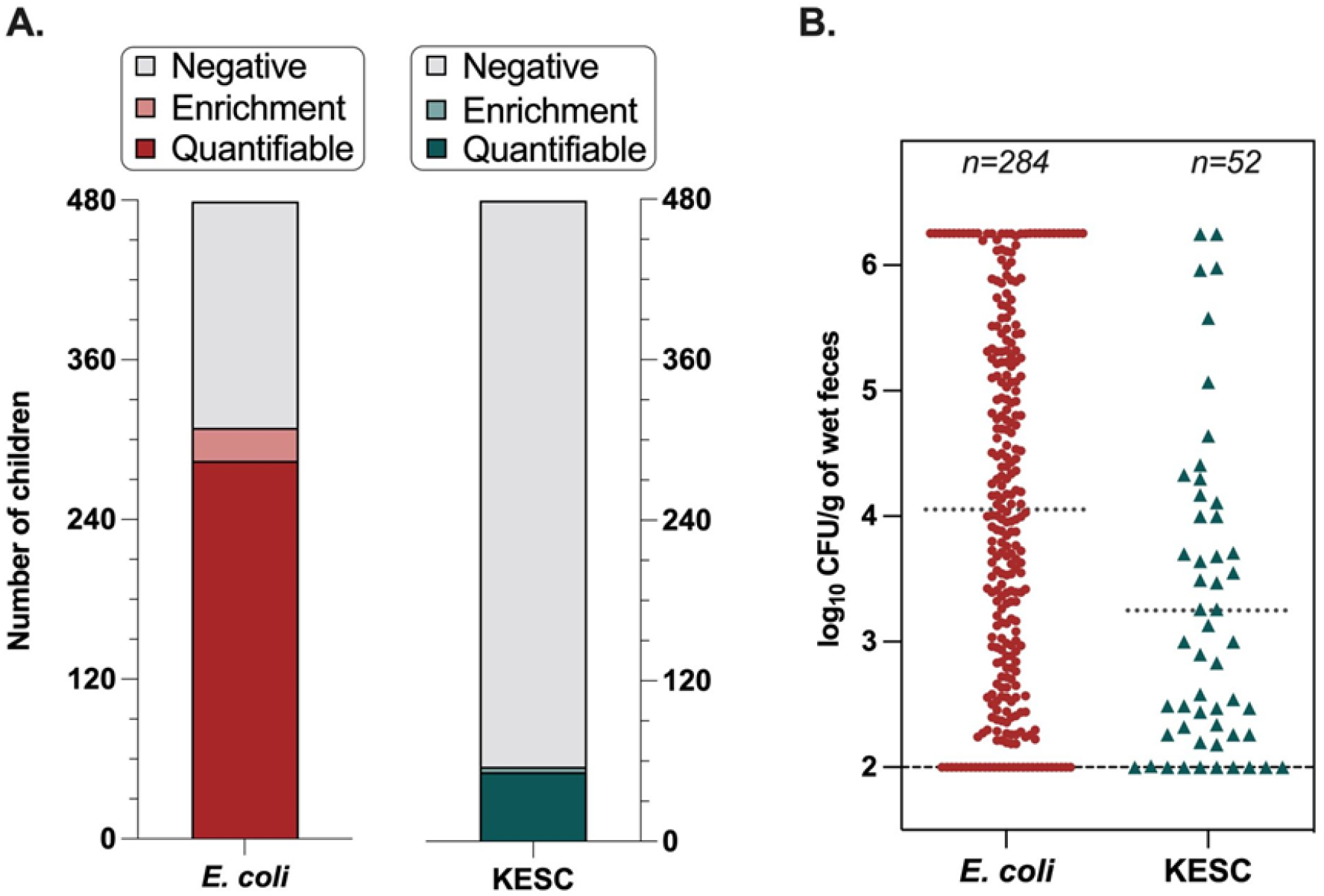
**A)** Fecal carriage rates of ESBL-*E. coli* (64.5%) and KESC (11.7%) in Bangladeshi children (n=479). Negative was assigned to samples where no growth was observed after direct plating onto the selective ESBL agar or after the pre-enrichment step; Enrichment corresponds to samples with presumptive ESBL-*E. coli* or -KESC colonies after the pre-enrichment step; Quantifiable corresponds to samples with presumptive ESBL-*E. coli* or -KESC colonies after direct plating onto the selective ESBL agar. **B)** Concentrations of ESBL-*E. coli* (4.05 log_10_ CFU/g-wet feces (± SD 1.42)) and -KESC (3.25 log_10_ CFU/g-wet feces (± SD 1.23)) amongst samples with direct quantitative positive cultures.

Based on both presence of colorless colonies, including light brown colonies indicative of species of *Proteus*, *Providencia*, and *Moraganella,* on the agar plates and the sample distribution relative to the lower LOD, these results likely underestimate the prevalence and concentration of ESBL-E within the population. We detected colorless colonies in 23% (n = 111) of fecal samples, of which 4% (n = 19) were negative for both KESC and ß-glucuronidase-producing *E. coli*. The colorless colonies could also include *E. coli* strains that do not possess the ß-glucuronidase enzyme, so a subset may be ESBL-producing *E. coli* strains.^47^ Additionally, fitting normal distributions to the log_10_-transformed ESBL-*E. coli* and ESBL-KESC concentrations (*fitdistcens* package in R) suggest population carriage is describable by mean (± SD) concentrations of 2.6 (±2.5) and 1.8 (± 3.0) log_10_ CFU/g-wet, respectively (Fig. 2). Based on these distributions, an estimated additional 21% of children (overall prevalence of 86%) were likely carriers of ESBL-*E. coli* and an additional 15% of children (overall prevalence of 37%) were likely carriers of ESBL-KESC at concentrations between 0 log_10_ CFU/g-wet and the lower LOD of 1.7 log_10_ CFU/g-wet.

**Figure 2.**
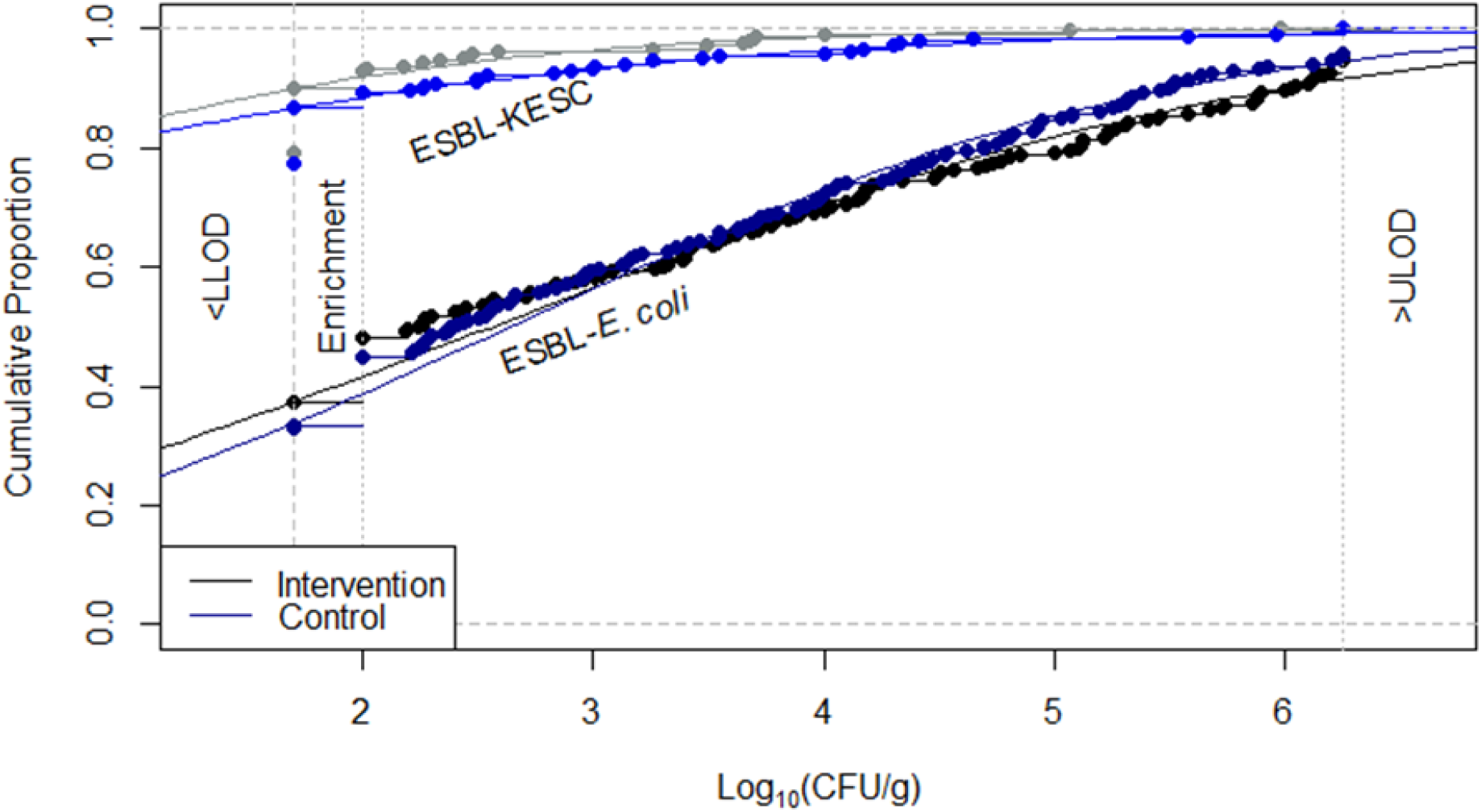
Empirical and theoretical normal cumulative distribution functions of concentrations of ESBL-*E. coli* and -KESC (log_10_ CFU/g-wet feces) amongst Bangladeshi children (n = 479) within the control and intervention groups

In a subset of 113 samples in which ESBL-*E. coli* concentrations were quantifiable, we also estimated total *E. coli* mean concentrations (± SD) of 5.6 (± 1.5) log_10_ CFU/g-wet. *E. coli* concentrations did not vary significantly between treatment and control (Welch two sample t-test, t = 0.65, df = 110, p = 0.51) or study site (t = 0.28, df = 74, p = 0.78). Among the 113 samples in the subset, ESBL-*E. coli* comprised the median [interquartile range] 4% [0.5%, 29%] of all *E. coli* (Fig. 3). However, in 15 (13%) samples, ESBL *E. coli* concentrations were higher than total *E. coli* concentrations, suggesting the variability of the method at the quantification level and/or the possibility that CHROMagar Orientation underestimates total *E. coli* concentrations (Fig. 3). Again, there was no difference in the proportion of ESBL *E. coli* between treatment and control (Wilcox rank sum test, w = 1757, p = 0.34) or study site (w = 1492, p = 0.93).

**Figure 3.**
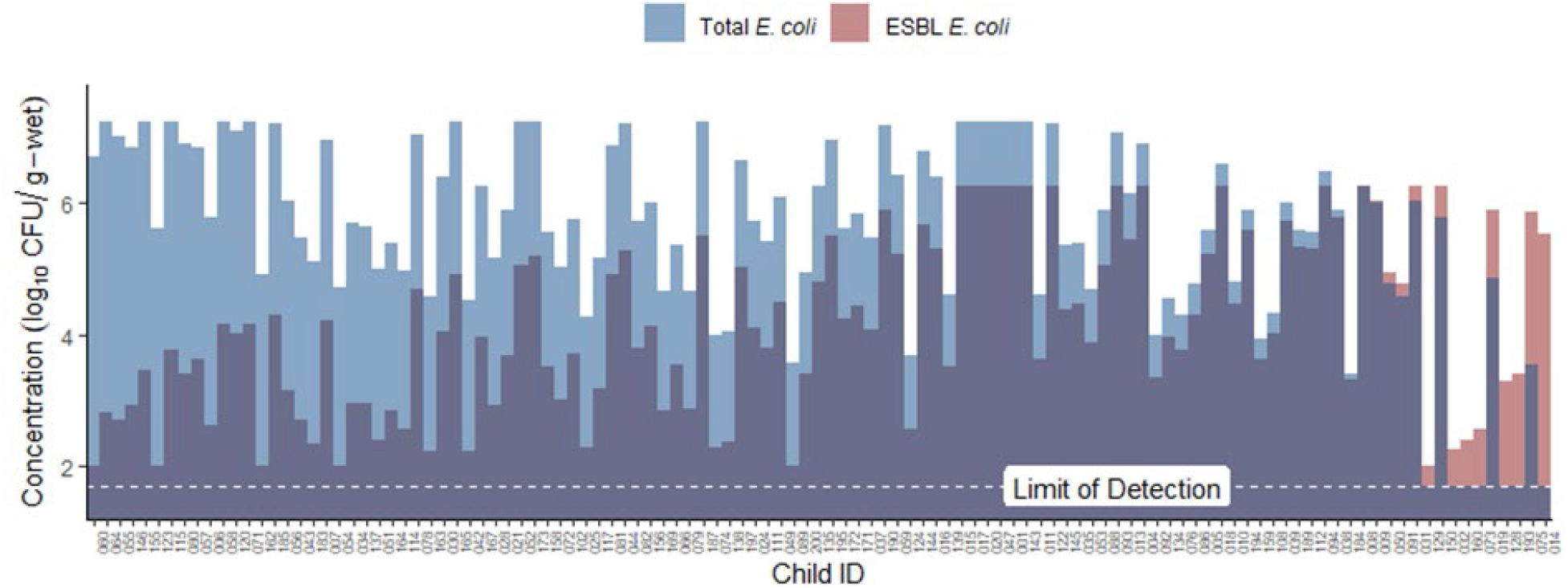
ESBL-*E. coli* fecal concentrations and corresponding total E. coli as measured on CHROM orientation agar in a subset of 113 samples. In 15 (13%) of the samples, total *E. coli*i concentrations were lower than ESBL-*E. coli*. The lower limit of detection (LLOD) for both ESBL-*E. coli* and total *E. coli* is 1.7 log_10_ CFU/g-wet.

### Impact of water chlorination on ESBL-E fecal carriage

Fecal carriage rates and fecal concentrations of ESBL-*E. coli* and ESBL-KESC were not different between the intervention group and the active control group (Fig. 4, Table 2, Table 3). Specifically, carriage rates of ESBL-*E. coli* was and 4.0% (67% vs. 63%) and for ESBL-KESC was 3.4% (13% vs. 10%) higher in the control group than in the intervention group, but the difference was not statistically significant when controlling for study site and age. Corresponding relative risks [95% CI] of the intervention for ESBL-*E. coli* was 0.98 [0.78, 1.23] and for ESBL-KESC was 0.76 [0.44, 1.29] (Table 2). Additionally, concentrations of ESBL-*E. coli* were 0.07 log_10_ CFU/g-wet lower in the control group (3.15 [2.94, 3.36] vs. 3.08 [2.89, 3.27]), and concentrations of ESBL-KESC were 0.10 log_10_ CFU/g-wet higher in the control group (1.92 [1.83, 2.02] vs. 1.82 [1,76, 1.88]), but neither relationship was statistically significant when controlling for study site and age (Fig. 4, Table 3).

**Figure 4.**
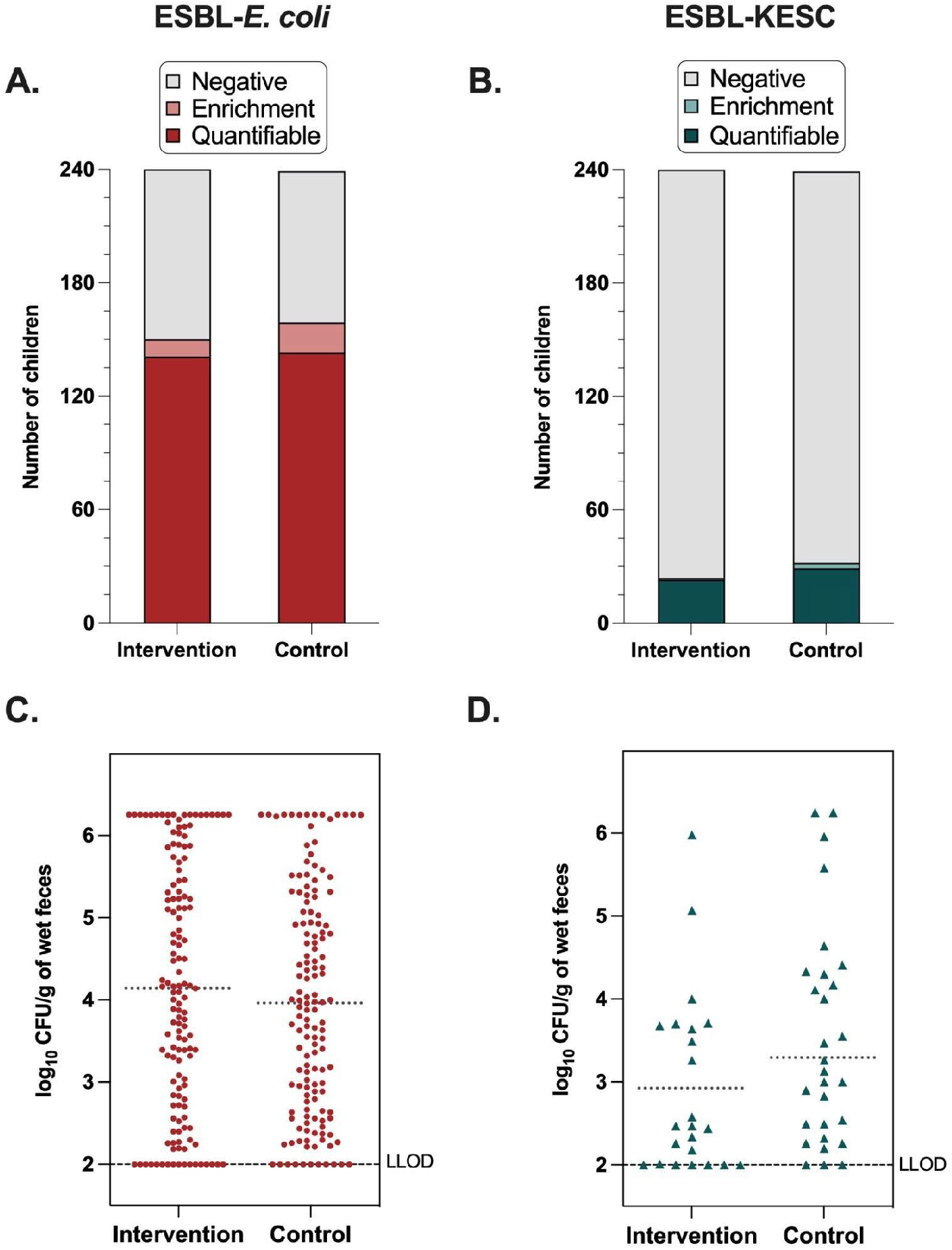
Fecal carriage rates of **A**) ESBL-*E. coli* and **B)** ESBL-KESC in the intervention and control groups. Negative was assigned to samples where no growth was observed after direct plating onto the CHROMID ESBL agar or after the enrichment step; Enrichment corresponds to samples with presumptive ESBL-*E. coli* colonies after the enrichment step; Quantifiable corresponds to samples with presumptive ESBL-*E. coli* colonies after direct plating onto the CHROMID ESBL agar. Concentrations of **C**) ESBL-*E. coli* and **D**) ESBL-KESC in the intervention and control groups amongst samples with direct positive cultures (quantifiable). The dotted horizontal line is the mean log_10_ CFU/g-wet feces in the intervention and control groups; the LLOD is indicated.

**Table 2.**
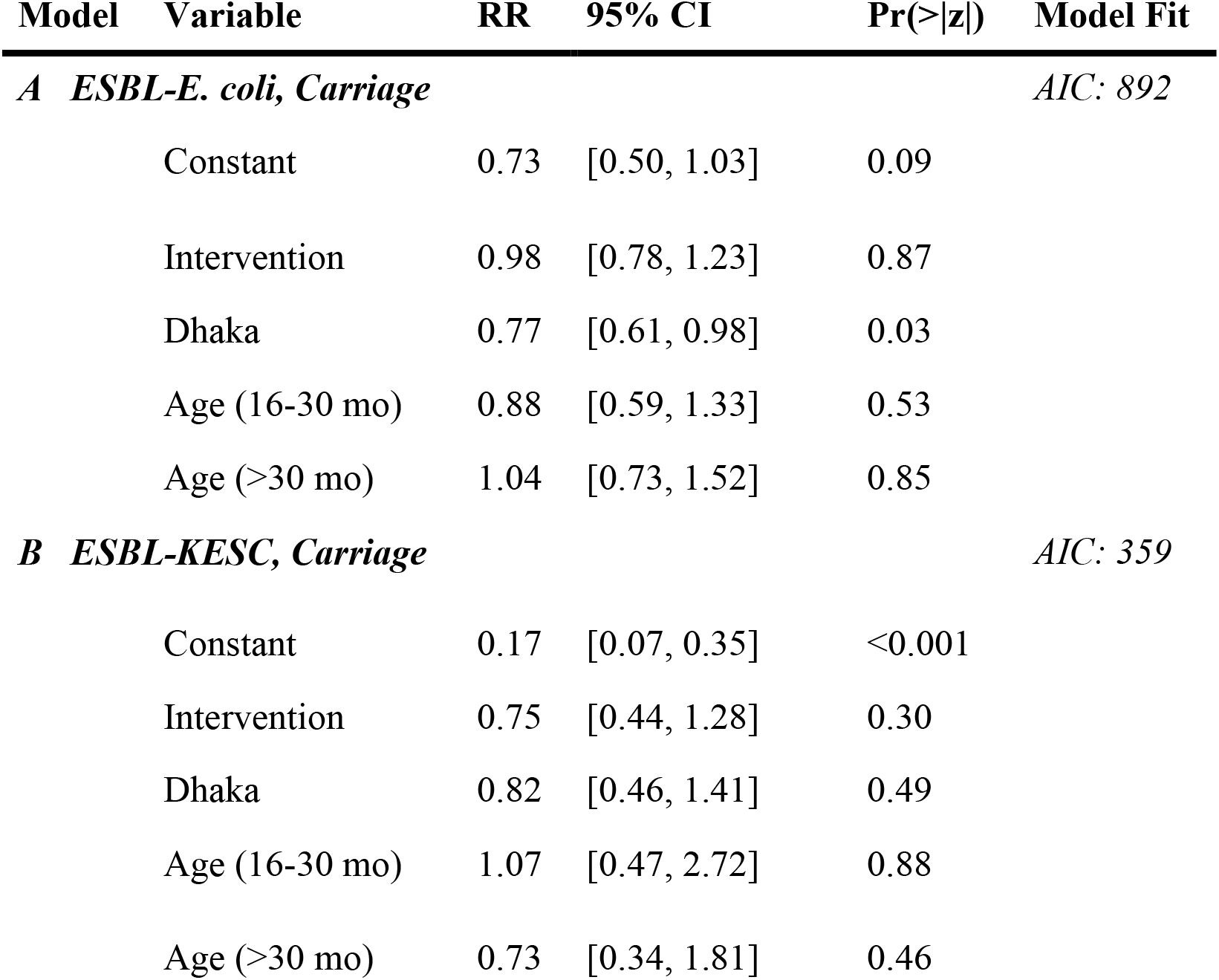
Impact of the drinking water chlorination intervention on children’s carriage of ESBL-*E. coli* and ESBL-KESC controlling for study site and age, as determined using modified Poisson regression. RR is Relative Risk and CI is the 95% Confidence Interval. Statistical significance is defined at α = 0.05.

| Model | Variable | RR | 95% CI | Pr(>\|z\|) | Model Fit |
| --- | --- | --- | --- | --- | --- |
| **A ESBL-*E. coli*, Carriage** |  |  |  |  | *AIC: 892* |
|  | Constant | 0.73 | [0.50, 1.03] | 0.09 |  |
|  | Intervention | 0.98 | [0.78, 1.23] | 0.87 |  |
|  | Dhaka | 0.77 | [0.61, 0.98] | 0.03 |  |
|  | Age (16-30 mo) | 0.88 | [0.59, 1.33] | 0.53 |  |
|  | Age (>30 mo) | 1.04 | [0.73, 1.52] | 0.85 |  |
| **B ESBL-KESC, Carriage** |  |  |  |  | *AIC: 359* |
|  | Constant | 0.17 | [0.07, 0.35] | <0.001 |  |
|  | Intervention | 0.75 | [0.44, 1.28] | 0.30 |  |
|  | Dhaka | 0.82 | [0.46, 1.41] | 0.49 |  |
|  | Age (16-30 mo) | 1.07 | [0.47, 2.72] | 0.88 |  |
|  | Age (>30 mo) | 0.73 | [0.34, 1.81] | 0.46 |  |

**Table 3.**
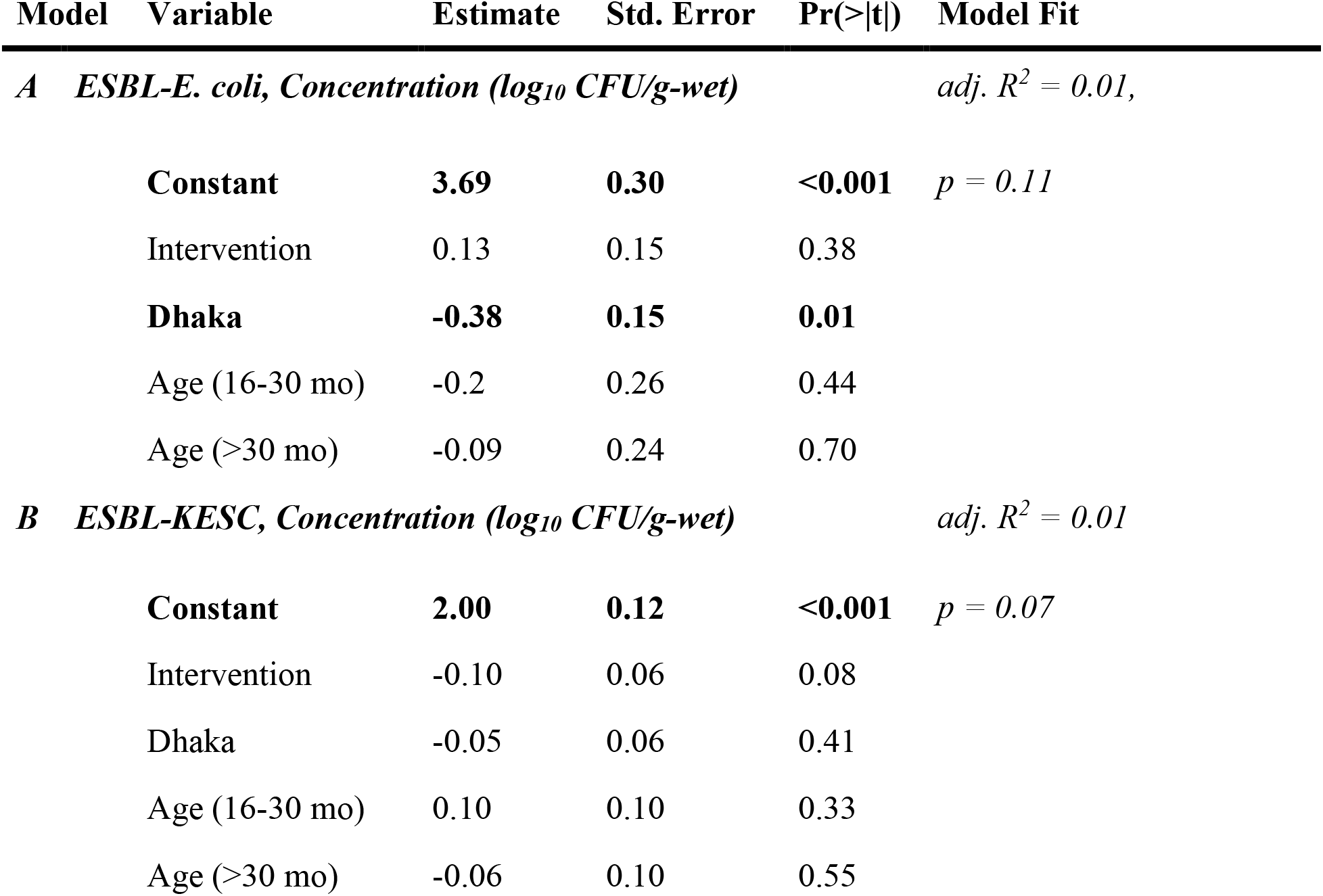
Impact of the drinking water chlorination intervention on concentration of ESBL-*E. coli* and ESBL-KESC in children’s feces controlling for study site and age, as determined using multiple linear regression. Emboldened variables are statistically significant defined at α = 0.05.

| Model | Variable | Estimate | Std. Error | Pr(>\|t\|) | Model Fit |
| --- | --- | --- | --- | --- | --- |
| **A ESBL-*E. coli*, Concentration ($\log_{10}$ CFU/g-wet)** |  |  |  |  | *adj. R*<sup>2</sup> = 0.01, |
|  | **Constant** | **3.69** | **0.30** | **<0.001** | *p* = 0.11 |
|  | Intervention | 0.13 | 0.15 | 0.38 |  |
|  | **Dhaka** | **-0.38** | **0.15** | **0.01** |  |
|  | Age (16-30 mo) | -0.2 | 0.26 | 0.44 |  |
|  | Age (>30 mo) | -0.09 | 0.24 | 0.70 |  |
| **B ESBL-KESC, Concentration ($\log_{10}$ CFU/g-wet)** |  |  |  |  | *adj. R*<sup>2</sup> = 0.01 |
|  | **Constant** | **2.00** | **0.12** | **<0.001** | *p* = 0.07 |
|  | Intervention | -0.10 | 0.06 | 0.08 |  |
|  | Dhaka | -0.05 | 0.06 | 0.41 |  |
|  | Age (16-30 mo) | 0.10 | 0.10 | 0.33 |  |
|  | Age (>30 mo) | -0.06 | 0.10 | 0.55 |  |

### Risk factors for ESBL-E. coli fecal carriage

ESBL-*E. coli* carriage was associated with study site and the number of households in a compound, but not with gender, age, caregiver reported antibiotic use in the last two months, visit to a treatment facility in the last two months, duration of study enrollment, or number of people in a household (Table 4). In Tongi, the carriage rate was 16.7 percentage points (71.3% to 54.6%) higher than in Dhaka and mean concentration of ESBL-*E. coli* was 0.36 log_10_ CFU/g-wet stool higher than in Dhaka. The number of households within a compound was inversely related to both percent carriage rate and mean concentrations of ESBL-*E. coli* (Table 4). Controlling for the study site did not influence the effect size or statistical significance of any of the risk factors with the exception of the number of households within a compound. Notably, the number of households within a compound is associated with study site (Table 1).

**Table 4.**
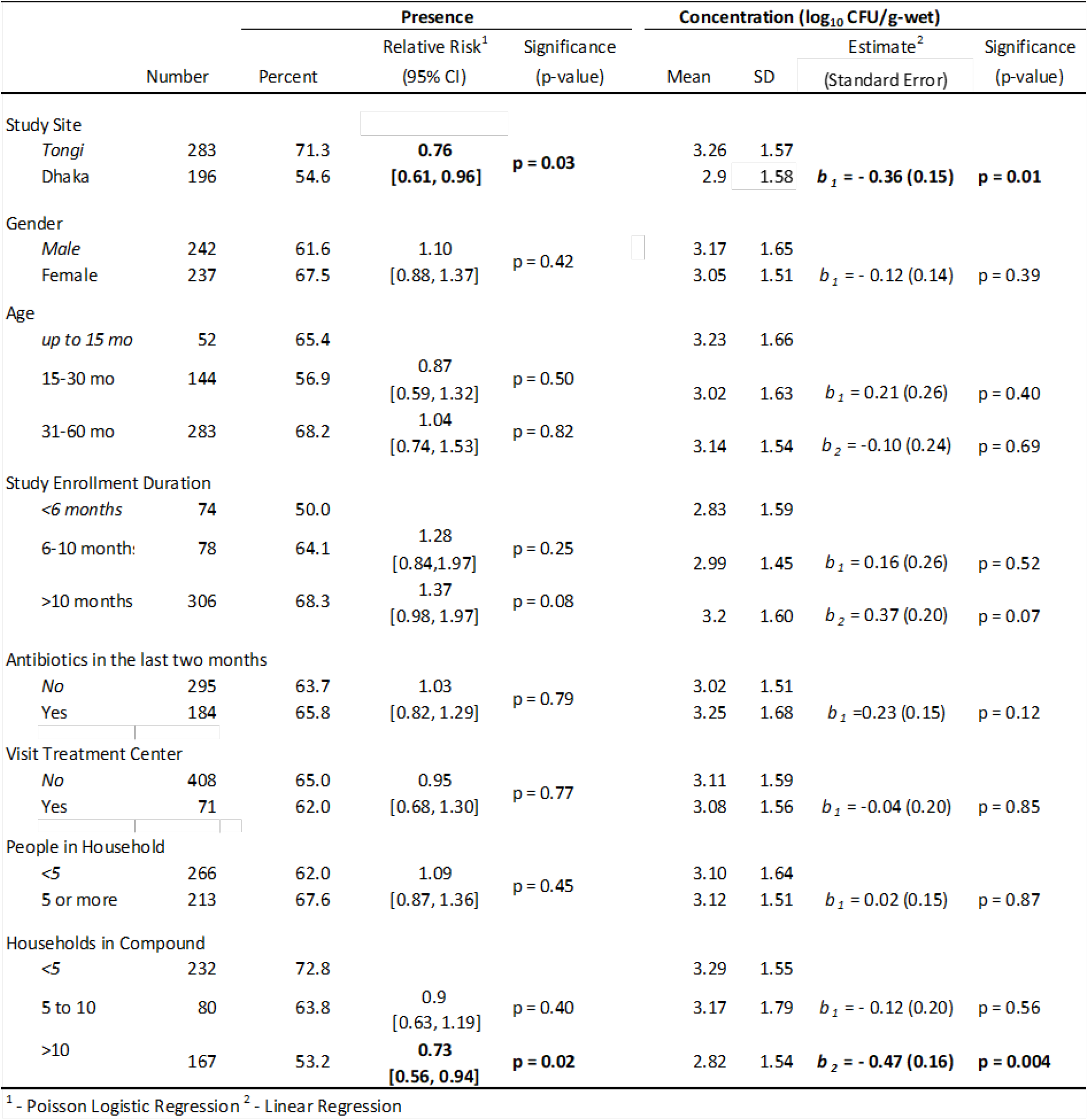
Impact of additional risk factors on children’s carriage and fecal concentration of ESBL-*E. coli*. Mean and standard deviation of concentrations are calculated using the lower LOD (1.7 log_10_ CFU / g-wet) for samples with no detectable ESBL-*E. coli*. Impact of risk factors on carriage determined using odds ratios derived from Poisson regression, and impact on concentration determined using effect size estimates derived from linear regression. Reference levels are italicized. For risk factors with statistical significance (p-value) < 0.05, odds ratios, estimates, and p-values are emboldened.

ESBL-KESC carriage was associated with antibiotic use in the past two months, but not with study site, child gender, recent visit to a treatment facility, duration of study enrollment, the number of people in the household, or the number of households in the compound (Table 5). Specifically, the percentage of children with ESBL-KESC was almost double amongst those who reported antibiotic use in the last two months compared to those that did not (relative risk of 1.99, p = 0.01). Mean concentration of ESBL-KESC was 0.21 log_10_ CFU/g-wet higher amongst children who reported antibiotic use in the past two months compared to those that did not (p < 0.001, Table 5).

**Table 5.** Impact of additional risk factors on children’s carriage and fecal concentration of ESBL-KESC. Mean and standard deviation of concentrations are calculated using the lower LOD (1.7 log_10_ CFU / g-wet) for samples with no detectable ESBL-KESC. Impact of risk factors on carriage determined using odds ratios derived from Poisson regression, and the impact on concentration is determined using effect size estimates derived from linear regression. Reference levels are italicized. For risk factors with statistical significance (p-value) < 0.05, odds ratios and estimates are emboldened.

| | Number | Percent | Presence | | Concentration ($\log_{10}$ CFU/g-wet) | | | |
| --- | --- | --- | --- | --- | --- | --- | --- | --- |
|  |  |  | Relative Risk<sup>1</sup><br>(95% CI) | Significance<br>Pr(>\|z\|) | Mean | SD | Estimate<sup>2</sup><br>(Standard Error) | Significance<br>(p-value) |
| Study Site |  |  |  |  |  |  |  |  |
| Tongi | 283 | 12.7 | 0.80 | p = 0.43 | 1.89 | 0.68 |  |  |
| Dhaka | 196 | 10.2 | [0.46, 1.37] | | 1.84 | 0.55 | $b_1 = -0.06$ (0.06) | p = 0.32 |
| Gender |  |  |  |  |  |  |  |  |
| Male | 242 | 11.2 | 1.10 | p = 0.73 | 1.84 | 0.56 |  |  |
| Female | 237 | 12.2 | [0.65, 1.86] | | 1.9 | 0.69 | $b_1 = 0.06$ (0.06) | p = 0.29 |
| Age |  |  |  |  |  |  |  |  |
| up to 15 mo | 52 | 13.4 |  |  | 1.87 | 0.52 |  |  |
| 15-30 mo | 144 | 13.9 | 1.03<br>[0.46, 2.63] | p = 0.94 | 1.96 | 0.85 | $b_1 = 0.09$ (0.10) | p = 0.37 |
| 31-60 mo | 283 | 10.2 | 0.76<br>[0.35, 1.89] | p = 0.52 | 1.82 | 0.5 | $b_2 = -0.04$ (0.09) | p = 0.63 |
| Study Enrollment Duration |  |  |  |  |  |  |  |  |
| <6 months | 74 | 12.2 |  |  | 1.8 | 0.37 |  |  |
| 6-10 months | 78 | 6.4 | 0.53<br>[0.16, 1.53] | p = 0.25 | 1.79 | 0.39 | $b_1 = -0.02$ (0.10) | p = 0.88 |
| >10 months | 306 | 13.1 | 1.07<br>[0.55, 2.36] | p = 0.85 | 1.92 | 0.74 | $b_2 = 0.11$ (0.08) | p = 0.17 |
| Antibiotics in the last two months |  |  |  |  |  |  |  |  |
| No | 295 | 8.5 | 1.99 | p = 0.01 | 1.79 | 0.42 |  |  |
| Yes | 184 | 16.8 | [1.18, 3.39] | | 1.99 | 0.85 | $b_1 = 0.21$ (0.06) | p < 0.001 |
| Visit Treatment Center |  |  |  |  |  |  |  |  |
| No | 408 | 12.0 | 0.82 | p = 0.63 | 1.86 | 0.61 |  |  |
| Yes | 71 | 9.9 | [0.34, 1.70] | | 1.9 | 0.73 | $b_1 = 0.03$ (0.08) | p = 0.67 |
| People in Household |  |  |  |  |  |  |  |  |
| <5 | 266 | 11.3 | 1.08 | p = 0.77 | 1.86 | 0.58 |  |  |
| 5 or more | 213 | 12.2 | [0.64, 1.82] | | 1.88 | 0.68 | $b_1 = 0.03$ (0.06) | p = 0.64 |
| Households in Compound |  |  |  |  |  |  |  |  |
| <5 | 232 | 12.1 |  |  | 1.87 | 0.59 |  |  |
| 5 to 10 | 80 | 15.0 | 1.24<br>[0.61, 2.39] | p = 0.53 | 2.02 | 0.93 | $b_1 = 0.15$ (0.08) | p = 0.06 |
| >10 | 167 | 9.6 | 0.79<br>[0.42, 1.45] | p = 0.46 | 1.81 | 0.48 | $b_2 = -0.06$ (0.06) | p = 0.37 |

<sup>1</sup> - Poisson Logistic Regression <sup>2</sup> - Linear Regression

Because the impact of the intervention on diarrhea was larger in Dhaka than in Tongi ^25^, subgroup analysis was conducted to investigate intervention impacts on the fecal carriage rates and fecal concentrations of ESBL-*E. coli* and ESBL-KESC within Dhaka and Tongi separately. The impacts of the intervention within each study site were consistent with the primary analysis for ESBL-*E. coli* carriage and both ESBL-*E. coli* and ESBL-KESC fecal concentrations, when controlling for child age (Tables 6 and 7). For ESBL-KESC carriage, however, the intervention impact was larger in Dhaka (relative risk [95% CI] of 0.57 [0.23, 1.38]) than in Tongi (0.89 [0.45, 1.71]), though the intervention impact remained non significant despite the observed large effect size (Table 6).

**Table 6.** Subgroup analysis by study site (Dhaka, Tongi) on impact of the drinking water chlorination intervention on children’s carriage of ESBL-*E. coli* and ESBL-KESC controlling for age, as determined using modified Poisson regression. RR is Relative Risk and CI is the 95% Confidence Interval. Statistical significance is defined at α = 0.05.

| Model | Variable | RR | 95% CI | Pr(>\|z\|) | Model Fit |
| --- | --- | --- | --- | --- | --- |
| **A** | ***ESBL-E. coli, Carriage (Dhaka)*** |  |  |  | *AIC: 351* |
|  | Constant | 0.54 | [0.27, 0.98] | 0.06 |  |
|  | Intervention | 0.95 | [0.65, 1.40] | 0.79 |  |
|  | Age (16-30 mo) | 0.95 | [0.49, 1.98] | 0.88 |  |
|  | Age (>30 mo) | 1.09 | [0.60, 2.19] | 0.78 |  |
| **B** | ***ESBL-KESC, Carriage (Dhaka)*** |  |  |  | *AIC: 137* |
|  | **Constant** | **0.13** | **[0.02, 0.45]** | **<0.007** |  |
|  | Intervention | 0.57 | [0.23, 1.38] | 0.21 |  |
|  | Age (16-30 mo) | 1.28 | [0.32, 8.47] | 0.76 |  |
|  | Age (>30 mo) | 0.89 | [0.23, 5.79] | 0.88 |  |
| **C** | ***ESBL-E. coli, Carriage (Tongi)*** |  |  |  | *AIC: 547* |
|  | Constant | 0.74 | [0.47, 1.11] | 0.17 |  |
|  | Intervention | 1.00 | [0.76, 1.32] | 0.99 |  |
|  | Age (16-30 mo) | 0.84 | [0.52, 1.41] | 0.49 |  |
|  | Age (>30 mo) | 1.00 | [0.66, 1.61] | 0.97 |  |
| **D** | ***ESBL-KESC, Carriage (Tongi)*** |  |  |  | *AIC: 227* |
|  | **Constant** | **0.17** | **[0.06, 0.39]** | **<0.001** |  |
|  | Intervention | 0.89 | [0.45, 1.71] | 0.72 |  |
|  | Age (16-30 mo) | 0.97 | [0.36, 3.04] | 0.95 |  |
|  | Age (>30 mo) | 0.67 | [0.27, 2.01] | 0.42 |  |

Analysis of the water chlorination intervention impact on the subgroup of 306 (64%) of the children for whom enrollment duration was longer than ten months showed no substantial difference in impact compared to that observed for the entire cohort (Tables 2, 3 and 8). Notably, this analysis shifted the average age of the children from 32 to 46 months.

### Beta-lactamase encoding genes in children’s fecal metagenomes

We included data from 95 samples (n = 49 control, n = 46 treatment) in our analysis of beta-lactamase encoding genes in fecal metagenomes with an average of 38.9 million 150 bp paired end reads per sample. The most common beta-lactamase-encoding genes (*bla*) detected were *cfxA* (n = 89 out of 95, or 94%), *bla*_ACI_ (75%), *bla*_TEM_ (72%), and *bla*_OXA_ (60%) (Table 9, Fig. 5). There was no significant difference in the relative abundance or occurrence of any *bla* gene or allele (Table 9). Only one gene allele, *bla*_CTX-M-15_, was significantly influenced by the study site (estimate = -4.1, SE = 0.91, p = <0.001), as it was detected significantly less frequently in Tongi than in Dhaka (Table 9, Fig. 5). Carbapenemase-encoding genes were not detected among children’s feces in this subset.

**Figure 5.**
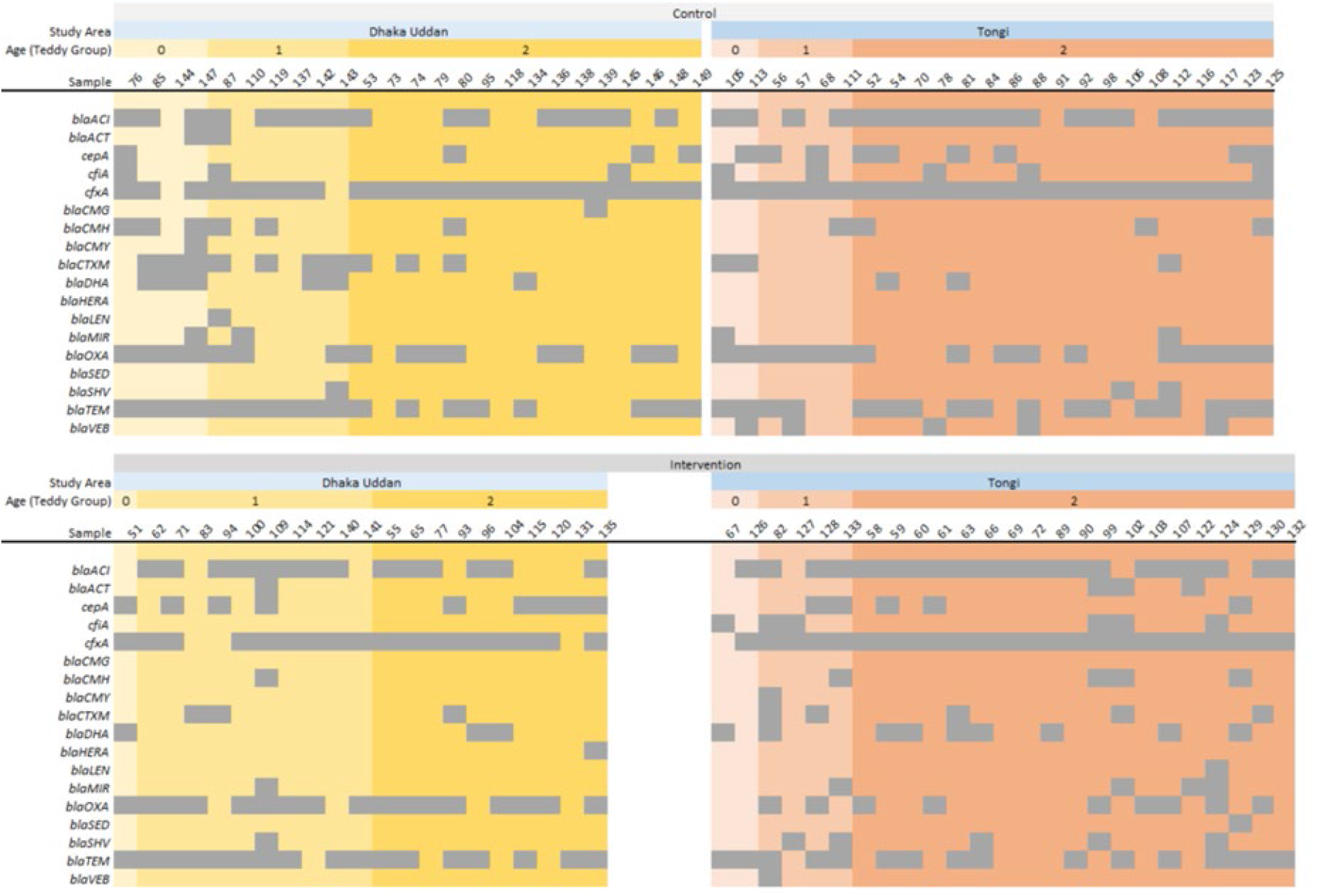
Beta-lactamase encoding genes in fecal samples (n = 95) of Bangladeshi children detected using metagenomics were not statistically significantly different between the control (n = 49) and the intervention (n = 46). Grey indicates the presence of the genes.

**Table 7.** Subgroup analysis by study site (Dhaka, Tongi) on impact of the drinking water chlorination intervention on concentration of ESBL-*E. coli* and ESBL-KESC in children’s feces controlling for study site and age, as determined using multiple linear regression. Emboldened variables are statistically significant defined at α = 0.05.

| Model | Variable | Estimate | Std. Error | Pr(>\|t\|) | Model Fit |
| --- | --- | --- | --- | --- | --- |
| **A** *ESBL-E. coli, Concentration (log<sub>10</sub> CFU/g-wet) (Dhaka)* |  |  |  |  | *adj. R<sup>2</sup> = -0.02, p = 0.99* |
|  | **Constant** | **2.86** | **0.38** | **<0.001** |  |
|  | Intervention | 0.05 | 0.23 | 0.83 |  |
|  | Age (16-30 mo) | 0.03 | 0.40 | 0.94 |  |
|  | Age (>30 mo) | -0.01 | 0.38 | 0.97 |  |
| **B** *ESBL-KESC, Concentration (log<sub>10</sub> CFU/g-wet) (Dhaka)* |  |  |  |  | *adj. R<sup>2</sup> = 0.01, p = 0.20* |
|  | **Constant** | **1.94** | **0.13** | **<0.001** |  |
|  | Intervention | -0.11 | 0.08 | 0.17 |  |
|  | Age (16-30 mo) | 0.04 | 0.14 | 0.77 |  |
|  | Age (>30 mo) | -0.10 | 0.13 | 0.45 |  |
| **C** *ESBL-E. coli, Concentration (log<sub>10</sub> CFU/g-wet) (Tongi)* |  |  |  |  | *adj. R<sup>2</sup> = 0.00, p = 0.46* |
|  | **Constant** | **3.37** | **0.30** | **<0.001** |  |
|  | Intervention | 0.19 | 0.19 | 0.31 |  |
|  | Age (16-30 mo) | -0.37 | 0.33 | 0.26 |  |
|  | Age (>30 mo) | -0.15 | 0.31 | 0.63 |  |
| **D** *ESBL-KESC, Concentration (log<sub>10</sub> CFU/g-wet) (Tongi)* |  |  |  |  | *adj. R<sup>2</sup> = 0.01, p = 0.21* |
|  | **Constant** | **1.92** | **0.13** | **<0.001** |  |
|  | Intervention | -0.10 | 0.08 | 0.22 |  |
|  | Age (16-30 mo) | 0.13 | 0.14 | 0.33 |  |
|  | Age (>30 mo) | -0.03 | 0.13 | 0.82 |  |

**Table 8.** Study enrollment duration subgroup analyses for the children (n = 306, 64%) exposed to the drinking water chlorination intervention for 10 or more months. Model outcomes are the primary and secondary analyses investigating impact of intervention on children’s carriage and fecal concentration of ESBL-E. coli and ESBL-KESC, controlling for study site and child age, as determined using modified Poisson regression (Models A and B) and multiple linear regression (Models C and D). RR is Relative Risk and CI is the 95% Confidence Interval. Statistical significance is defined at α = 0.05. For risk factors with statistical significance (p-value) < 0.05, odds ratios and estimates are emboldened.

| *Target* | *Variable* | *RR* | *[95% CI]* | *Pr(>\|z\|)* | *AIC* |
| --- | --- | --- | --- | --- | --- |
| A. ESBL-E. coli, carriage |  |  |  |  | 580 |
|  | **Intercept** | **0.68** | **[0.51, 0.88]** | **0.005** |  |
|  | Intervention | 1.04 | [0.79, 1.36] | 0.75 |  |
|  | Dhaka | 0.82 | [0.60, 1.10] | 0.20 |  |
|  | Age (>30 mo) | 1.09 | [0.83, 1.45] | 0.55 |  |
| B. ESBL-KESC, carriage |  |  |  |  | 246 |
|  | **Intercept** | **0.22** | **[0.12, 0.36]** | **<0.001** |  |
|  | Intervention | 0.74 | [0.39, 1.38] | 0.34 |  |
|  | Dhaka | 0.80 | [0.38, 1.55] | 0.52 |  |
|  | Age (>30 mo) | 0.56 | [0.30, 1.04] | 0.07 |  |
| **Model** | **Variable** | **Estimate** | **Std. Error** | **Pr(>\|t\|)** | **Model Fit** |
| C. ESBL-E. coli, Concentration (log<sub>10</sub> CFU/g-wet) |  |  |  |  | adj. R<sup>2</sup> = 0.01 |
|  | **Constant** | **3.23** | **0.18** | **<0.001** | p = 0.09 |
|  | Intervention | 0.33 | 0.18 | 0.08 |  |
|  | Dhaka | -0.36 | 0.20 | 0.06 |  |
|  | Age (>30 mo) | -0.11 | 0.19 | 0.55 |  |
| D. ESBL-KESC, Concentration (log<sub>10</sub> CFU/g-wet) |  |  |  |  | adj. R<sup>2</sup> = 0.01 |
|  | **Constant** | **2.09** | **0.08** | **<0.001** | p = 0.10 |
|  | Intervention | -0.12 | 0.08 | 0.18 |  |
|  | Dhaka | -0.03 | 0.09 | 0.72 |  |
|  | **Age (>30 mo)** | **-0.18** | **0.09** | **0.03** |  |

**Table 9.**
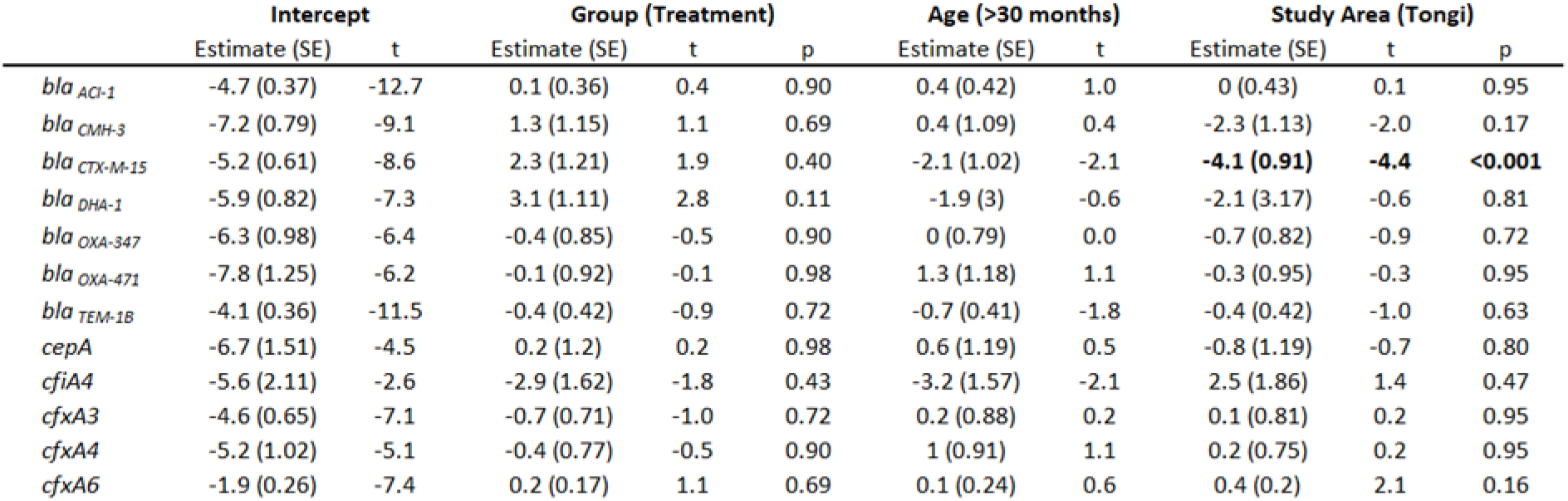
Logistic regression results for the impact of the treatment relative to the control on detection of beta-lactamase gene alleles. SE is the standard error of the estimate, and p-values are corrected for false discovery rate using Benjamini-Hochberg method.

Of the 95 samples analyzed, 29 (31%) were from children who did not have culturable ESBL-E. Metagenomics revealed one or more *bla* genes in all of these children. For example, ESBL or other *bla* genes commonly found in *Enterobacteriaceae* were also detected in the metagenomics data from children without culturable ESBL-E, including *bla*_CTX-M_ in 6 (21%)*, bla*_OXA_ in 16 (55%), and *bla*_TEM_ in 21of the samples (72%). In addition, other *bla*-encoding genes, commonly found in members of the intestinal microflora, were detected in children without culturable ESBL bacteria. For example, *bla*_ACI_ was detected in 28 (78%) samples.

### Genomics analyses of fecal ESBL-E. coli isolates

The 86 *E. coli* isolates representing one isolate per child were diverse and covered the eight *E. coli* phylogroups described in the literature, with the most frequent phylogroups, B1 (23.3%), B2 (22.1%), A (20.9%), and D (20.9%), showing similar prevalence (Figs. 6, 7, 8). Phylogroups A and B1, known to contain generalistic strains isolated from a variety of hosts and secondary environments, were found more frequently in children in the intervention group compared to the control group (Fig. 7). Conversely, the phylogroups B2 and D, known to contain the majority of extraintestinal pathogenic strains, were more common in children from the control group compared to the intervention group (Fig. 7). However, there were no significant differences in phylogroup distribution between control and intervention groups (Pearson’s Chi-squared test, χ^2^ = 8.6, p = 0.28). The phylogroup distribution was also similar across the two sites Dhaka and Tongi (Pearson’s Chi-squared test, χ^2^ = 10.6, p = 0.16).

**Figure 6.**
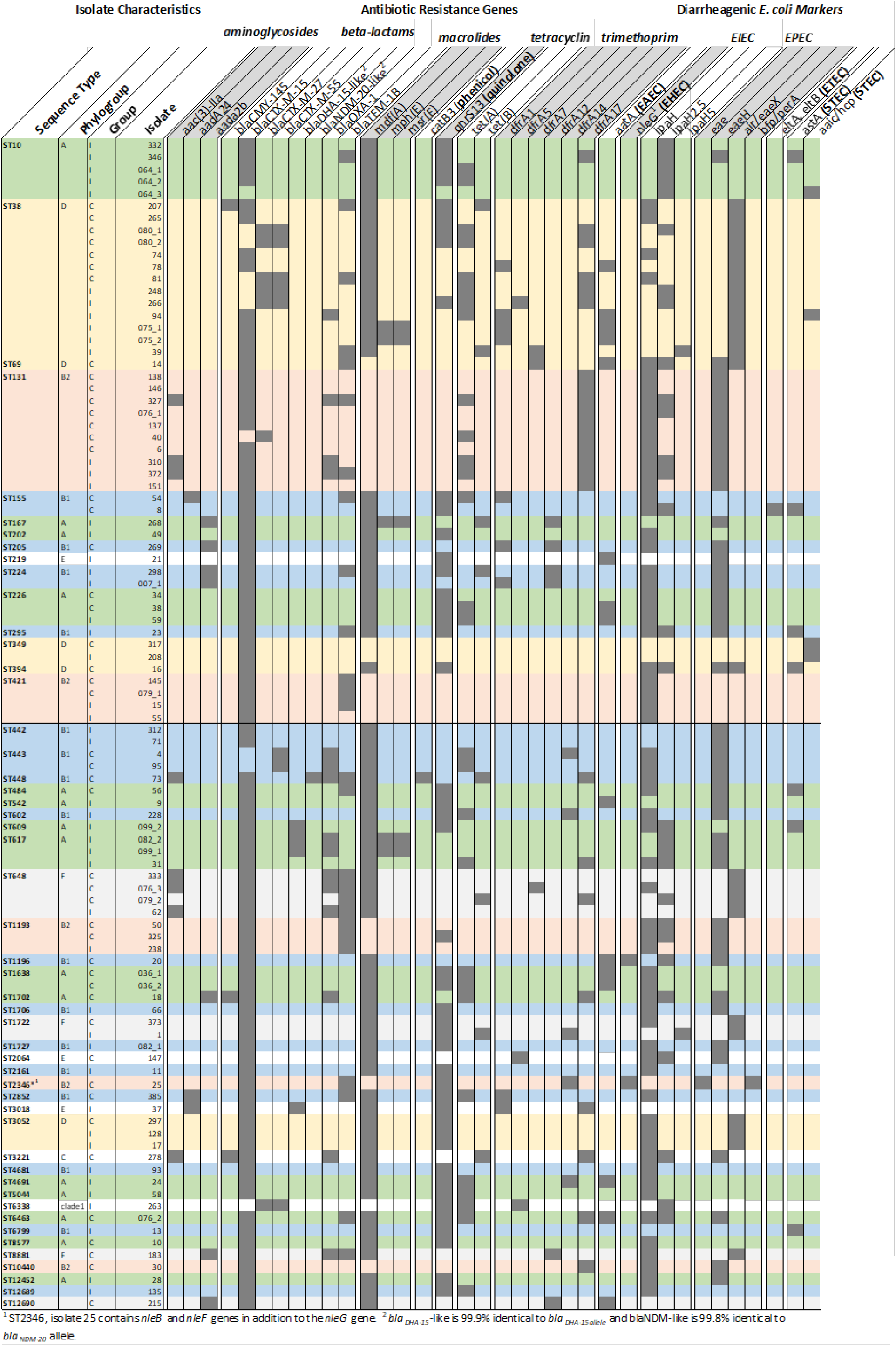
*E. coli* isolate characteristics. Child ID/ Isolate with succeeding “_1” represents isolates from samples from which additional isolates were also sequenced (see Figure 9). Background color corresponds to the phylogroup color-coding scheme used in Figure 8.

**Figure 7.**
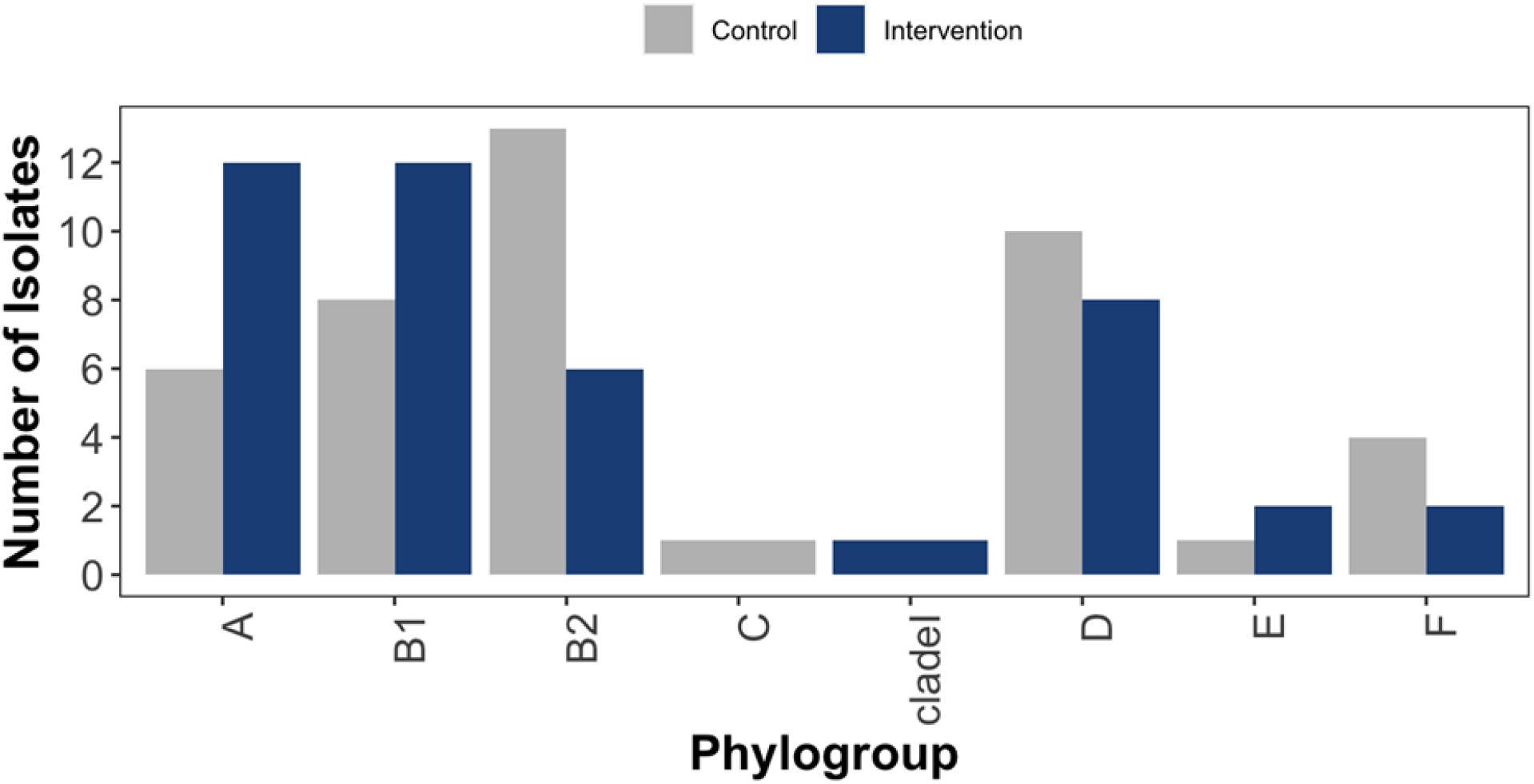
Distribution of *E. coli* phylogroups (n = 86) from Bangladeshi children in the Control and Intervention groups.

**Figure 8.**
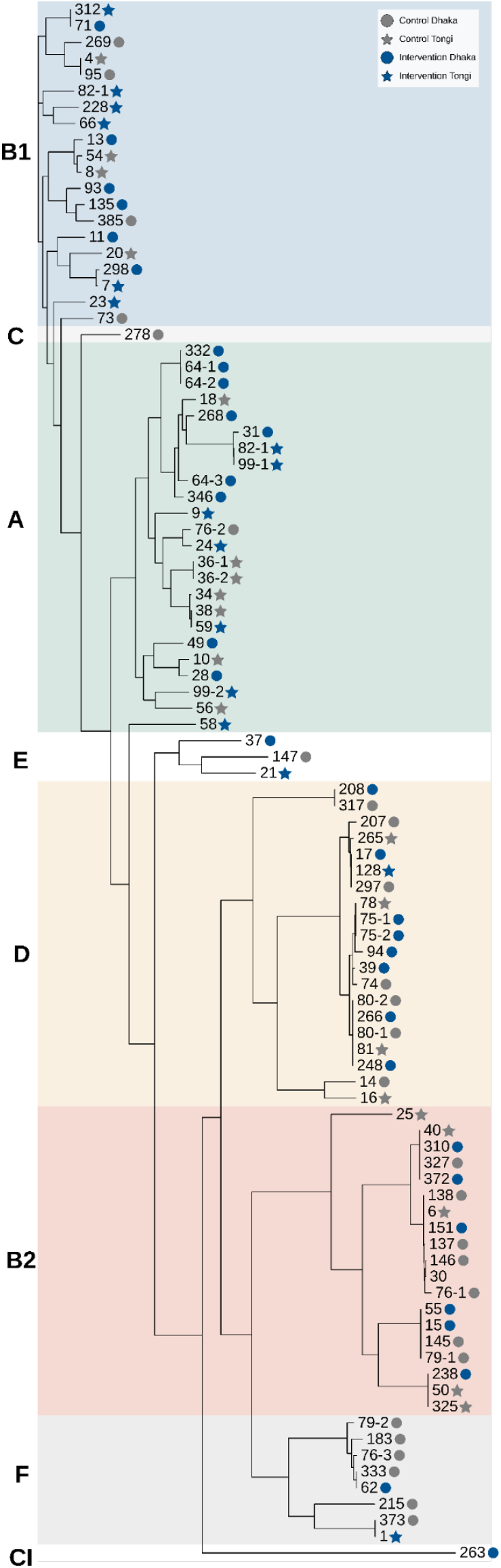
Phylogenetic tree of 96 fecal ESBL-*E. coli* isolates from 86 children, representing 1, 2 or 3 isolates per child. Numbers corresponding to children from the control (n = 43) and intervention (n = 43) are labeled grey and blue, respectively. Study site is denoted by shape, with Dhaka as circles and Tongi as stars. Phylogroup is denoted by background color.

With the sequenced *E. coli*, we identified 50 unique sequence types (STs, Fig.S5). The most prevalent sequence types encountered were ST38 (12.8%) and ST131 (11.6%), lineages of widespread distribution associated with multidrug resistance and extraintestinal infections (mainly urinary tract and bloodstream infections).^48^ Sequence types ST167 (1 isolate) and ST224 (2 isolates) as well as antimicrobial resistance genes *bla*_CTX-M-15_, *bla*_NDM-type_, *bla*_CMY-type_, *aac6’-lb-cr, qnrS* were previously found in clinical *E. coli* isolates in Bangladesh.^49–51^

Antibiotic resistance genes were widespread in the isolates. As expected, given that we cultured *E. coli* on an agar selective for ESBL-producing Enterobacteriaceae, almost all (99%) of *E. coli* carried an ESBL gene, with *bla*_CTX-M-15_ as the most prevalent (90%). Almost half of the isolates (42%) contained multiple beta-lactamase genes, typically *bla*_CTX-M-15_ co-occurring with *bla*_TEM-1_ and/or *bla*_OXA-1_. Genes conferring resistance to macrolides (73%), quinolones (48%), tetracyclines (37%), and trimethoprim (48%) were also common (Fig. 6). Comparing the *E. coli* isolates based on the presence of any antibiotic resistance gene, there was no significant difference between the isolates from the control group relative to the isolates in the intervention group, when controlling for the false discovery rate (Table 10). Notably, *bla*_CTX-M-15_ was detected in 90% of *E. coli* isolates but only 22% of metagenomic samples (Figs. 5, 6), suggesting differential sensitivity of culture-based as compared to metagenomic methods.

**Table 10.** Frequency of antimicrobial-resistant and diarrheagenic gene detection within 86 sequenced *E. coli* fecal isolates from children in the control (n = 43) and intervention (n = 43) groups. Statistical significance of the difference in frequency was determined using Student t-test with p-value adjusted using Benjamini-Hochberg to account for multiple comparisons.

| Gene | Control | Intervention | p(adj) |
| --- | --- | --- | --- |
| aminoglycosides |  |  |  |
| *aac(3)-IIa* | 9% | 9% | 1.00 |
| *aadA24* | 5% | 2% | 0.88 |
| *aada2b* | 9% | 7% | 0.90 |
| beta-lactams |  |  |  |
| *bla*<sub>CMY-145</sub> | 7% | 2% | 0.72 |
| *bla*<sub>CTX-M-15</sub> | 88% | 91% | 0.90 |
| *bla*<sub>CTX-M-27</sub> | 7% | 7% | 1.00 |
| *bla*<sub>CTX-M-55</sub> | 9% | 7% | 0.90 |
| *bla*<sub>DHA-15-like</sub> | 0% | 5% | 0.62 |
| *bla*<sub>NDM-20-like</sub> | 2% | 0% | 0.72 |
| *bla*<sub>OXA-1</sub> | 19% | 12% | 0.79 |
| *bla*<sub>TEM-1B</sub> | 35% | 21% | 0.62 |
| macrolides |  |  |  |
| *mdf(A)* | 65% | 81% | 0.62 |
| *mph(E)* | 0% | 7% | 0.62 |
| *msr(E)* | 0% | 7% | 0.62 |
| phenicols |  |  |  |
| *catB3* | 2% | 0% | 0.72 |
| quinolone |  |  |  |
| *qnrS13* | 37% | 58% | 0.62 |
| tetracycline |  |  |  |
| *tet(A)* | 26% | 30% | 0.90 |
| *tet(B)* | 7% | 12% | 0.88 |
| trimethoprim |  |  |  |
| *dfrA1* | 9% | 7% | 0.90 |
| *dfrA5* | 2% | 5% | 0.88 |
| *dfrA7* | 2% | 2% | 1.00 |
| *dfrA12* | 7% | 7% | 1.00 |
| *dfrA14* | 5% | 7% | 0.90 |
| *dfrA17* | 30% | 19% | 0.72 |
| diarrheagenic *E. coli* genes |  |  |  |
| *aatA* | 14% | 14% | 1.00 |
| *nleG* | 5% | 0% | 0.62 |
| *ipaH* | 84% | 54% | 0.08 |
| *ipaH2.5* | 28% | 26% | 0.97 |
| *ipaH5* | 0% | 5% | 0.62 |
| *eae* | 2% | 0% | 0.72 |
| *eaeH* | 54% | 47% | 0.88 |
| *air/eaeX* | 28% | 21% | 0.88 |
| *bfp/perA* | 2% | 0% | 0.72 |
| *eltA, eltB* | 2% | 0% | 0.72 |
| *astA* | 7% | 7% | 1.00 |
| *aaic/hcp* | 2% | 5% | 0.88 |

Of the 86 *E. coli* isolates, 72% harbored genes conferring resistance to at least one agent in three or more antibiotic categories, with 79% (of 43 isolates) in the intervention compared to 65% (of 43 isolates) in the control (z = -1.45, p = 0.15).^52^ Nine (11%) isolates harbored genes conferring resistance to five or more antibiotic categories (Fig. 6). Antibiotic resistance genes typically shared 100% identity with known alleles. However, sample identity was lower for some reported alleles. For example, *mdf(A)* with reported identity of 98.1% was detected in 16 isolates, suggesting potential novel alleles..

*E. coli* genomes also contained genes specific to diarrheagenic *E. coli* (Fig. 6). The most prevalent were *ipaH* (66%) and *ipaH2.5* (30%) indicative of enteroinvasive *E. coli*; *eaeH* (47%) and *air/eaeX* (26%) indicative of enteropathogenic *E. coli;* and *astA* (7%) associated with enteroaggregative *E. coli,* enteropathogenic *E. coli,* enterotoxigenic *E. coli,* and Shiga toxin-producing *E. coli* (Table 10).^53–56^ Most diarrheagenic *E. coli* genes (*nleG, ipaH, ipaH2.5, eae, eaeH, air/eaeX, bfp/perA, eltA, eltB*) were detected more frequently in the control group than in the intervention group (Table 10). Nevertheless, there was no significant difference for any gene when controlling for multiple comparisons (Table 10).

For eight randomly selected children, more than one *E. coli* colony was sequenced (Fig. 9). In three children we were able to detect distinct sequence types. In four children, isolates shared the same sequence type and with <60 pairwise single nucleotide polymorphisms between core genomes (isolate 36-1 with 36-2, 64-1 with 64-2, 75-1 with 75-2, and 80-1 with 80-2, Fig.2 and Fig. 9). For context, pairwise single nucleotide polymorphism differences in *E. coli* of <40 have been used to define clones,^57^ and <100 to define clades.^58^ In two children, isolates from the same sequence type had large (>5000) pairwise single nucleotide polymorphisms (99-1 with 99-2 and 64-1/64-2 with 64-3). In one of the isolates (64-3), no ESBL gene was detected, despite having been cultured on selective CHROMID^®^ ESBL.

**Figure 9.**
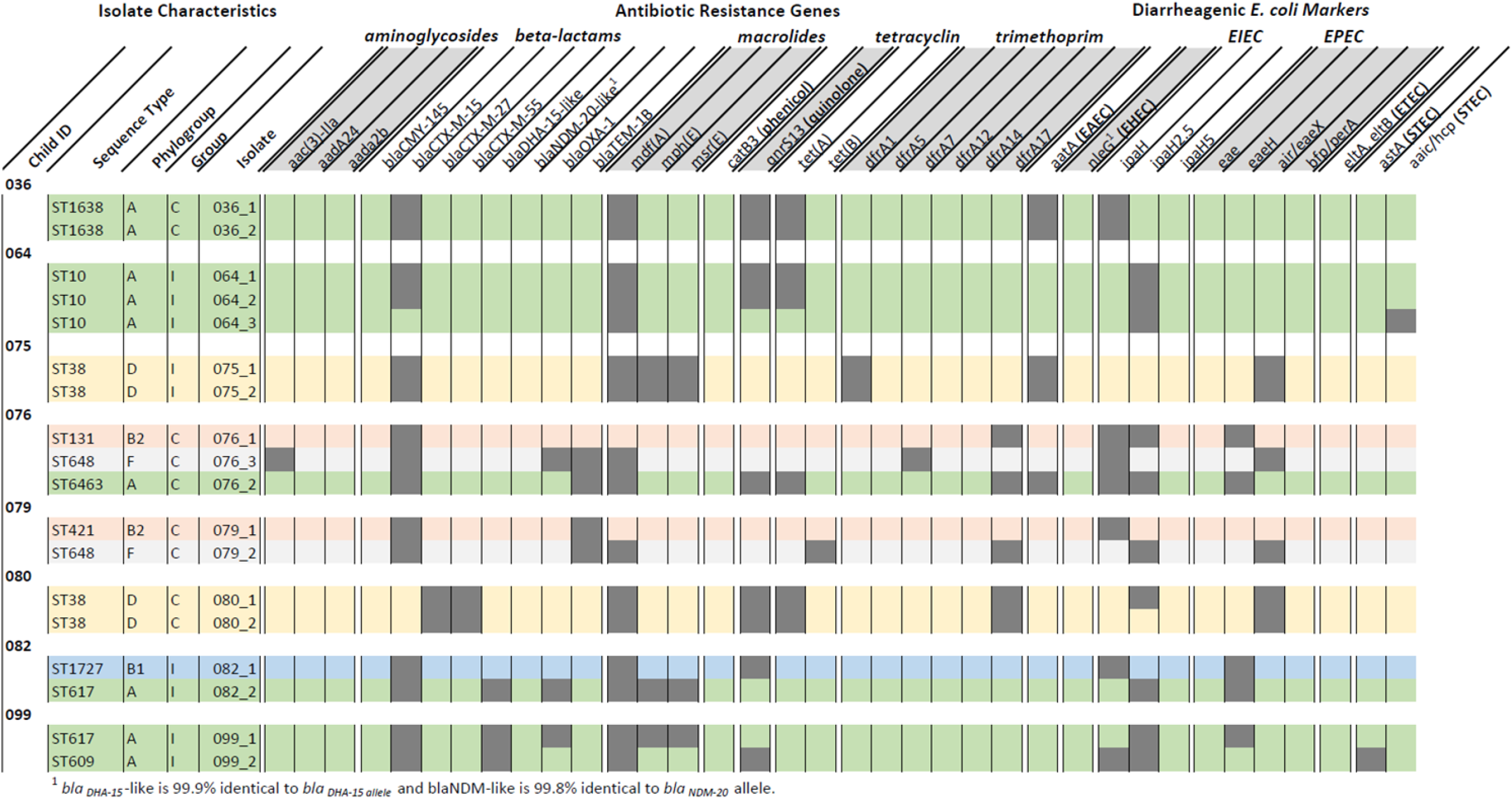
Multiple (two or three) *E. coli* isolates were sequenced from a subset of eight child fecal samples. Amongst half of the children (child IDs 076, 079, 082, and 099), we detected different strains, while for the remaining half (036, 064, 075, 080), isolates were clonal.

## DISCUSSION

Using a cluster-randomized controlled trial study design, we found that in-line drinking water chlorination did not significantly reduce fecal carriage or concentrations of ESBL-*E. coli* or ESBL-KESC in Bangladeshi children. Previous work demonstrated that in-line drinking water chlorination effectively reduced diarrheal disease and antibiotic use amongst Bangladeshi children.^25^ Reductions in diarrheal disease and antibiotic use are both posited to influence AMR carriage, which motivated our hypothesis that receiving chlorinated water could reduce fecal carriage rates of ESBL-E in Bangladeshi children.^15^ Further, metagenomic analysis identified no significant difference in the presence or relative abundance of beta-lactamase-encoding genes, and whole-genome sequencing of ESBL-*E. coli* isolates revealed only limited impact on the mechanisms of resistance. The findings highlight that in-line chlorination of drinking water, an example of an effective WASH intervention for controlling enteric pathogens via a single exposure route, was insufficient alone to influence ESBL*-E. coli* carriage. The high prevalence of ESBL-*E. coli* in the community and the multiple environmental transmission routes in this setting may have overwhelmed the impact of an intervention focused on water quality improvement alone. Given extensive support for WASH investments to combat AMR transmission in national and international action plans^14,15,17,59,60^, there is a clear need to identify conditions under which interventions will be effective.

### The study highlights high carriage rates of ESBL-*E. coli* amongst Bangladeshi children

Within this study, estimated prevalence of ESBL-*E. coli* in children was 65%, and may nevertheless be an underestimate. The estimate is influenced by the limit of detection of the culture method used in the study (≥ 10 CFU/g feces), and it is possible that a large proportion of Bangladeshi children with no detectable culturable ESBL-E were carriers at concentrations below our detection limit. Indeed, ESBL-encoding genes - including those commonly carried by *E. coli* - were detected in metagenomics data from some Bangladeshi children who tested negative for culturable ESBL-E. High prevalence could be explained by rapid acquisition and/or long periods of carriage after acquisition, two phenomena previously described for ESBL-*E. coli*.^61–63^ Rapid acquisition of ESBL-*E. coli* in Bangladesh is further supported by findings of high prevalence in both infants under one year old and adults, and evidence of acquisition in adults during travel to South Asia generally.^64–67^ ESBL-*E. coli* carriership is also long: In a study of adults in the Netherlands, time to loss of carriership was 1.1 years ^63^.

### The lack of a significant effect of drinking water chlorination on ESBL-*E. coli* carriage stands in contrast to the impact that the same intervention had on diarrheal disease

One explanation may be the higher prevalence of ESBL-*E. coli* in the community relative to enteric pathogens. Diarrheal prevalence, a crude indicator of enteric pathogen prevalence, was substantially lower in this cohort (diarrheal prevalence of 7-9%, depending on definition)^25^. Higher prevalence rates offer increased opportunities for transmission, potentially reducing intervention efficacy targeting one exposure route. Indeed, the comparative analysis of ESBL-KESC carriage supports this: ESBL-KESC was carried by a substantially lower portion of the children than ESBL-*E. coli* (12% compared to 65%), and the intervention was associated with a more pronounced reduction in ESBL-KESC (relative risk [95% CI]: 0.75 [0.44, 1.28] compared to 0.98 [0.78, 1.23] for ESBL-*E. coli*). Additionally, prior antibiotic use was statistically significantly associated with increased ESBL-KESC, whereas it had no impact on ESBL-*E. coli* carriage.

### A lack of an observed impact of water chlorination on ESBL-E carriage may also be attributed to the longer duration of ESBL-*E. coli* carriership relative to enteric pathogens

In contrast to the aforementioned median estimate of 1.1 years to loss of carriership in the Netherlands, enteric pathogen shedding duration is typically less than 30 days ^63,68^. Interventions such as drinking water treatment which interrupt exposures but do not affect carriage may not impact prevalence until there has been sufficient loss of carriage. In this study, the samples evaluated originated from an open cohort design in which median duration of exposure to the chlorine intervention was 10.7 months. This exposure period may be sufficient to observe effects of reduced exposure to enteric pathogens, but not to ESBL-*E. coli* given longer periods of carriage. However, this study found no evidence that ESBL-*E. coli* or ESBL-KESC fecal carriage or concentrations were impacted by the duration of the children’s exposure to the drinking water intervention up to the year assessed here. Future studies may consider monitoring intervention impacts targeting reductions in AMR exposures beyond time periods sufficient to monitor diarrheal disease (for example, longer than one year).

### Drinking water may also be a more important route of transmission for chlorine sensitive enteric pathogens than for ESBL-*E. coli*

The reported reduction in diarrheal disease by the intervention in Pickering et al. (2019) indicates that improvements to drinking water quality reduce enteric disease transmissi^on.25^ In contrast, the same intervention did not reduce the prevalence of carriage of antimicrobial resistance genes. Given the importance of multiple sources of environmental contamination in transmission of enteric pathogens, it is likely environmental exposures similarly contribute to ESBL-*E. coli* transmission.^68,69^ Estimated concentrations of ESBL-*E. coli* in fecal samples ranged from 10^2^ to more than 10^6^ CFU/g-wet feces, and accounted for an estimated median of 3.5% of all *E. coli* in the gut, suggesting transmissibility through the fecal-oral route. ESBL-*E. coli* is also transmitted via animals, with observed high rates of ESBL-*E. coli* colonization of backyard poultry in Bangladesh.^67,70^ Further, the widespread detection of ESBL-*E. coli* in soils, on hands, and in source and stored drinking water in Bangladesh supports the potential of its environmental transmission.^8,9,71–76^ ESBL-*E. coli* have also been identified in surface water ^77^ and wastewater in urban areas ^78,79^, posing risks to people - including children - who interact with open drains or are exposed to water from sewage overflows.^80^ Environmental interventions targeting a single route of exposure, even one that is responsible for a substantial portion of enteric pathogen transmission, may be insufficient to combat AMR transmission in regions of high AMR prevalence and multiple concurrent exposure routes. Improved understanding of AMR exposure routes, inclusive interfaces among animals, people, and the environment (“One Health”), may elucidate potential for multimodal interventions beyond in-line chlorination alone to reduce ESBL-*E. coli* carriage.

### Geographic differences in ESBL-*E. coli* carriage rates highlight the influence of environmental exposures on health risks

ESBL-*E. coli* carriage was 17% higher in Tongi than in Dhaka, regions that are geographically close (less than 15 km) in a high population density country. Higher rates of ESBL-*E. coli* carriage were also observed in Tongi when considering only the control group (72% in Tongi compared to 56% in Dhaka). Geographic influence on carriage of AMR is well-documented.^6^ For example, international travel to areas with high prevalence rates of AMR is a major risk factor for subsequent carriage.^81^ The cause for differences in ESBL-*E. coli* prevalence between the two sites is unclear. High prevalence rates are often attributed to combinations of inappropriate antibiotic consumption; inadequate WASH access and coverage; and deficient healthcare quality and access.^16^ However, communities included in the study from Tongi and Dhaka are both low-income communities with similar reported income levels, antibiotic use, and WASH and healthcare access.^25^ Notable differences are that participants in Tongi were less likely to report drinking water treatment at taps, were more likely to perceive their water sources to be unsafe, had higher access to private taps relative to Dhaka, and a higher proportion of people living in multi-dwelling households.^25^ As Tongi is not part of the Dhaka city corporation, wastewater and sewage disposal are also under different management. Additionally, when the drinking water intervention impacts on diarrhea were analyzed by subgroups, significant reductions were observed in Dhaka but not in Tongi ^25^. This suggests that other environmental, social or behavioral factors may also play a role in determining local AMR patterns.

### The ESBL-*E. coli* isolates identified in children in Dhaka and Tongi are potential sources of extraintestinal infections

Two of the *E. coli* sequence types (ST38 and ST131) that we commonly detected are frequently associated with extraintestinal infections (ExPEC: extraintestinal pathogenic *E. coli*). ST38 isolates are frequently associated with urinary tract infections^82^, and here all isolates harbored resistance genes to beta-lactams and macrolides, with some isolates additionally positive for resistance genes to quinolones, tetracyclines, and/or trimethoprim. All ST38 isolates in this study, as the ST38 isolates reported enteroaggregative *E. coli* in Europe,^83^ contained the *air/eaeX* gene, a putative enteroaggregative immunoglobulin repeat protein, thus highlighting the potential of these isolates to cause enteric disease. ST131 is the predominant lineage of extraintestinal pathogenic *E. coli* worldwide and frequently harbors the CTX-M-15 ESBL gene as observed in the present study.^84–87^ Although ESBL carriage is not an indicator of acute health issues, it is nevertheless the most significant risk factor for ESBL-*E. coli* bacteremia.^3^ The high rates of carriage observed here and elsewhere^64^ suggests Bangladeshi children are at increased risk of future AMR infections. Children may also contribute to circulation of ESBL-E in households ^88,89^, suggesting increased risks for carriage among adults.

This study is subject to limitations that could be addressed in future research. An important limitation is study power. The desired sample size (n = 600) was not reached due to the aforementioned issues with sample collection and processing. Additionally, the sample size calculation underestimated the prevalence of ESBL-*E. coli* in the children by 37%. As a result, the study was powered to detect an actual effect size of 15% reduction in ESBL-*E. coli*. It is unclear what reduction in AMR carriage would meaningfully improve health, but for comparison, Woerther et al. (2013) estimated an annual increase rate for ESBL carriage in South Asia of 7.2% ^6^. Similarly, based on the sample size, the impact of the intervention on ESBL-KESC was powered to detect a reduction in prevalence of at least 5.5%, which would correspond to an almost halving in prevalence rate from 13% to at least 7%. For comparison, we observed a 3% reduction in prevalence from 13% to 10%, a meaningful effect size (relative risk of 0.75) if it could be shown to be attributable to the intervention. Future studies investigating WASH intervention impacts on antimicrobial resistance carriage should include larger sample sizes sufficient to observe meaningful effects, such as that observed ESBL-KESC. Although this would necessitate clinical sample collection and analysis from 1000s of children, such a study design could mirror trials investigating WASH impacts on enteropathogens ^90^.

Additional limitations of the study were the open enrollment study design and limited duration of the intervention. Children who entered the study in the intervention group after the study launch were exposed to the intervention for only a subset of the trial period. Indeed, 16% of the children were enrolled for less than 6 months before fecal sample collection and so did not receive the full treatment effects. Notably, these children were characterized by lower (though not significantly) ESBL-E. coli and ESBL-KESC carriage rates, which could be explained by either their birth into the study (lower colonization rates are expected at birth)^64^ or their movement into the study from regions with lower ESBL-E carriage. Additionally, as previously discussed, the treatment duration observed here may be insufficient to observe impacts of environmental interventions on AMR. Future studies of WASH impacts on antimicrobial resistance carriage should consider closed enrollment, including a population with sufficient sample size to allow attrition such that samples are only included from children receiving the full benefits of treatment.*Finally, there was an* imbalance in sample sizes between the two study sites (Dhaka and Tongi). The inclusion here of fewer samples from Dhaka (where the intervention was more effective) than Tongi may have further limited the power of the study to observe a significant impact ^25^. With ESBL-KESC, subgroup analysis revealed a more substantial reduction in Dhaka than in either Tongi or the entire cohort. This suggests future evaluations of WASH impacts on fecal carriage of antimicrobial resistance may have more success if they are restricted to regions in which the interventions are effective against diarrheal diseases. Importantly, the study findings are representative of the two low-income urban communities in Dhaka and Tongi and may not be generalizable to other locales including high-income or rural communities in Bangladesh.

Our study findings have important implications, including emphasizing the need to identify WASH strategies effective at reducing AMR carriage. Despite calls within national and global AMR action plans for WASH, empirical evidence on effective interventions is lacking. Here, we demonstrate that in-line drinking water chlorination, an effective intervention at reducing diarrheal disease, had no meaningful impact on ESBL*-E. coli* among children in an area of high ESBL-*E. coli* prevalence. In contrast, a meaningful but non-significant reduction in ESBL-KESC carriage was observed, suggesting WASH interventions may be more effective at reducing AMR carriage when there is lower prevalence. Conceptually, investments in WASH remain a practical recommendation to tackle the threat of AMR. Nevertheless, empirical evidence is needed to identify the conditions under which WASH investments impact AMR. Gathering such evidence requires: 1) defining meaningful reductions in AMR carriage, 2) identifying WASH interventions with the potential to achieve these reductions (such as those with proven efficacy against diarrheal disease), 3) allowing the community sufficient exposure to the intervention to allow loss of AMR carriage, which may be substantially longer than traditional WASH intervention trials, and 4) evaluating a sufficient sample size to determine the significance of a meaningful reduction. We encourage future investigators to carefully consider these criteria when designing future intervention trials aimed at curbing the spread and selection of AMR.

## Data Availability

*E. coli* genomes are archived at NCBI BioProject PRJNA705080: Water Chlorination impacts on *E. coli* diversity in Bangladeshi child feces. Metagenomes are archived as NCBI Bioproject PRJNA706606.

## Author Contributions

MCM, AJP, TRJ conceived and supervised the study; MCM, MAI, VFL, SPL, AJP, and TRJ developed the proposal, planned experiments, and secured the funding; JS, SS, SPL, AJP collected and provided samples; MCM, EEG, LT, LC, TN processed samples; MCM, TRJ, MLN analyzed and interpreted data; MCM, EEG, TRJ wrote the original draft; all authors contributed to writing the final draft.

## Funding Source Statement

This work was funded by the Thrasher Research Fund (#14205). The funder had no role in data collection, data analysis, data interpretation, or writing of this report. MLN was supported by NIH award KL2TR002545 and the Stuart B. Levy Center for Integrated Management of Antimicrobial Resistance at Tufts (Levy CIMAR), a collaboration of Tufts Medical Center and the Tufts University Office of the Vice Provost for Research (OVPR) Research and Scholarship Strategic Plan (RSSP).

## Supporting information

CONSORT 2010 Checklist

## Data Availability

All data produced in the present study are available upon reasonable request to the authors

## Notes

### Competing Interest Statement

The authors have declared no competing interest.

### Clinical Trial

NCT02606981

### Author Declarations

The study protocol for the original trial25 was approved by the International Centre for Diarrhoeal Diseases Research, Bangladesh (icddr,b) scientific and ethical review committees (protocol number 14022) and the human subjects institutional review board at Stanford University (protocol number 30456). Within the original trial, informed written consent was obtained from all study participants as well as the owners of the water points enrolled. Consent included biospecimen collection for future unplanned analyses.

