## Supplementary material for "Drinking water chlorination impact on fecal carriage of extended-spectrum beta-lactamase-producing *Enterobacteriaceae* in Bangladeshi children in a double-blind, cluster-randomized controlled trial": CONSORT 2010 Checklist

Table 1: CONSORT 2010 checklist of information to include when reporting a cluster randomised trial

| Section/Topic | Item No | Standard Checklist item | Extension for cluster designs | Page No/ Section * |
|---|---|---|---|---|
| **Title and abstract** | | | | |
| | 1a | Identification as a randomised trial in the title | Identification as a cluster randomised trial in the title | Title (Montealegre, Maria C., et al. (2021)) ; Title (Pickering, Amy J., et al. (2019)) |
| | 1b | Structured summary of trial design, methods, results, and conclusions (for specific guidance see CONSORT for abstracts)[i][ii] | See table 2 | Abstract (Montealegre, Maria C., et al. (2021)) Abstract (Pickering, Amy J., et al. (2019)) |
| **Introduction** | | | | |
| **Background and objectives** | 2a | Scientific background and explanation of rationale | Rationale for using a cluster design | Methods, Participants (Pickering, Amy J., et al. (2019)) Not updated for this study (Monteleagre et al.) as cluster design was performed in the scope of Pickering, et al. (2019) |
| | 2b | Specific objectives or hypotheses | Whether objectives pertain to the the cluster level, the individual participant level or both | Methods, Study design and fecal samples (Montealegre, Maria C., et al. (2021)), Introduction (Pickering, Amy J., et al. (2019)) |
| **Methods** | | | | |
| **Trial design** | 3a | Description of trial design (such as parallel, factorial) including allocation ratio | Definition of cluster and description of how the design features apply to the clusters | Methods, Study design and fecal samples (Montealegre, Maria C., et al. (2021)), Methods, Participants (Pickering, Amy J., et al. (2019)) |
| | 3b | Important changes to methods after trial commencement (such as eligibility criteria), with reasons | | **N/A** |
| **Participants** | 4a | Eligibility criteria for participants | Eligibility criteria for clusters | Methods, Study design and fecal samples (Montealegre, Maria C., et al. (2021)); Methods, Participants (Pickering, Amy J., et al. (2019)) |
| | 4b | Settings and locations where the data were collected | | Methods, Study design and fecal samples (Montealegre, Maria C., et al. (2021)); Methods, Study design (Pickering, Amy J., et al. (2019)) |
| | | The interventions for each group with sufficient details to | Whether interventions pertain to the | Methods, Study design and fecal samples (Montealegre, Maria C., et al. (2021)); |

| | | | | |
|---|---|---|---|---|
| **Interventions** | 5 | allow replication, including how and when they were actually administered | cluster level, the individual participant level or both | Methods, Intervention delivery (Pickering, Amy J., et al. (2019)) |
| **Outcomes** | 6a | Completely defined pre-specified primary and secondary outcome measures, including how and when they were assessed | Whether outcome measures pertain to the cluster level, the individual participant level or both | Methods, Study design and fecal samples (Montealegre, Maria C., et al. (2021)); Methods - outcomes (Pickering, Amy J., et al. (2019)) |
| | 6b | Any changes to trial outcomes after the trial commenced, with reasons | | Methods - outcomes (Pickering, Amy J., et al. (2019)) Not updated for this study (Monteleagre et al.) as any changes were reported in Pickering et al. (2019) |
| **Sample size** | 7a | How sample size was determined | Method of calculation, number of clusters(s) (and whether equal or unequal cluster sizes are assumed), cluster size, a coefficient of intracluster correlation (ICC or *k* ), and an indication of its uncertainty | Methods, Study design and fecal samples (Montealegre, Maria C., et al. (2021)); Methods – statistical analysis (Pickering, Amy J., et al. (2019)) |
| | 7b | When applicable, explanation of any interim analyses and stopping guidelines | | Procedures (Pickering, Amy J., et al. (2019)) Not updated for this study (Monteleagre et al.) |
| **Randomisation:** | | | | |
| **Sequence generation** | 8a | Method used to generate the random allocation sequence | | Methods, Randomisation (Pickering, Amy J., et al. (2019)) Not updated for this study (Monteleagre et al.) as random allocation was performed in the scope of Pickering, et al. (2019) |
| | 8b | Type of randomisation; details of any restriction (such as blocking and block size) | Details of stratification or matching if used | Methods, Randomisation (Pickering, Amy J., et al. (2019)) Not updated for this study (Monteleagre et al.) as randomisation was performed in the scope of Pickering, et al. (2019) |
| **Allocation concealment mechanism** | 9 | Mechanism used to implement the random allocation sequence (such as sequentially numbered containers), describing any steps taken to conceal the sequence until interventions were assigned | Specification that allocation was based on clusters rather than individuals and whether allocation concealment (if any) was at the cluster level, the individual participant level or both | Methods, Randomisation (Pickering, Amy J., et al. (2019)) Not updated for this study (Monteleagre et al.) as random allocation was performed in the scope of Pickering, et al. (2019) |

| | | | | |
|---|---|---|---|---|
| **Implementation** | 10 | Who generated the random allocation sequence, who enrolled participants, and who assigned participants to interventions | Replace by 10a, 10b and 10c | Methods, Randomisation (Pickering, Amy J., et al. (2019))<br><br>Not updated for this study (Monteleagre et al.) as random allocation was performed in the scope of Pickering, et al. (2019) |
| | 10a | | Who generated the random allocation sequence, who enrolled clusters, and who assigned clusters to interventions | Methods, Randomisation (Pickering, Amy J., et al. (2019))<br><br>Not updated for this study (Monteleagre et al.) as random allocation was performed in the scope of Pickering, et al. (2019) |
| | 10b | | Mechanism by which individual participants were included in clusters for the purposes of the trial (such as complete enumeration, random sampling) | Methods, Randomisation (Pickering, Amy J., et al. (2019))<br><br>Not updated for this study (Monteleagre et al.) as random allocation and enrollment were performed in the scope of Pickering, et al. (2019) |
| | 10c | | From whom consent was sought (representatives of the cluster, or individual cluster members, or both), and whether consent was sought before or after randomisation | Methods, Study design (Pickering, Amy J., et al. (2019))<br><br>Not updated for this study (Monteleagre et al.) as consent was sought in the scope of Pickering, et al. (2019) |
| **Blinding** | 11a | If done, who was blinded after assignment to interventions (for example, participants, care providers, those assessing outcomes) and how | | Methods, Study design and fecal samples (Montealegre, Maria C., et al. (2021));<br>Methods, intervention delivery and masking (Pickering, Amy J., et al. (2019)) |
| | 11b | If relevant, description of the similarity of interventions | | Methods, intervention delivery and masking (Pickering, Amy J., et al. (2019))<br>Not updated for this study (Monteleagre et al.) |
| **Statistical methods** | 12a | Statistical methods used to compare groups for primary and secondary outcomes | How clustering was taken into account | Methods, Statistical analyses (Montealegre, Maria C., et al. (2021));<br>Methods – statistical analysis (Pickering, Amy J., et al. (2019)) |
| | [illegible] | Methods for additional analyses, such as subgroup analyses | | Methods, Statistical analyses (Montealegre, Maria C., et al. (2021)); |

| | | | | |
|---|---|---|---|---|
| | 12b | [illegible] and adjusted analyses | | Methods – statistical analysis (Pickering, Amy J., et al. (2019)) |
| **Results** | | | | |
| **Participant flow (a diagram is strongly recommended)** | 13a | For each group, the numbers of participants who were randomly assigned, received intended treatment, and were analysed for the primary outcome | For each group, the numbers of clusters that were randomly assigned, received intended treatment, and were analysed for the primary outcome | Methods and Results (Montealegre, Maria C., et al. (2021));<br>Figure 1 (Pickering, Amy J., et al. (2019)) |
| | 13b | For each group, losses and exclusions after randomisation, together with reasons | For each group, losses and exclusions for both clusters and individual cluster members | Methods, Study design and fecal samples (Montealegre, Maria C., et al. (2021));<br>Figure 1 (Pickering, Amy J., et al. (2019)) |
| **Recruitment** | 14a | Dates defining the periods of recruitment and follow-up | | Results (Pickering, Amy J., et al. (2019))<br>Not updated for this study (Monteleagre et al.), as recruitment and follow-up were conducted in Pickering et al. (2019). |
| | 14b | Why the trial ended or was stopped | | Results and Discussion (Pickering, Amy J., et al. (2019))<br>Not updated for this study (Monteleagre et al.), as trial was ended in scope of Pickering et al. (2019). |
| **Baseline data** | 15 | A table showing baseline demographic and clinical characteristics for each group | Baseline characteristics for the individual and cluster levels as applicable for each group | Table 1 (Montealegre, Maria C., et al. (2021));<br>Table 1 (Pickering, Amy J., et al. (2019)) |
| **Numbers analysed** | 16 | For each group, number of participants (denominator) included in each analysis and whether the analysis was by original assigned groups | For each group, number of clusters included in each analysis | Results (Montealegre, Maria C., et al. (2021));<br>Figure 1 (Pickering, Amy J., et al. (2019)) |
| **Outcomes and estimation** | 17a | For each primary and secondary outcome, results for each group, and the estimated effect size and its precision (such as 95% confidence interval) | Results at the individual or cluster level as applicable and a coefficient of intracluster correlation (ICC or k) for each primary outcome | Results (Montealegre, Maria C., et al. (2021));<br>Figure 1, Table 3 (Pickering, Amy J., et al. (2019)) |
| | 17b | For binary outcomes, presentation of both absolute and relative effect sizes is recommended | | Results (Montealegre, Maria C., et al. (2021));<br>Table 3 (Pickering, Amy J., et al. (2019)) |
| **Ancillary analyses** | 18 | Results of any other analyses performed, including subgroup analyses and adjusted analyses, distinguishing pre-specified from exploratory | | Results (Montealegre, Maria C., et al. (2021));<br>Results, Table 2, Table 3 (Pickering, Amy J., et al. (2019)) |
| **Harms** | 19 | All important harms or unintended effects in each group (for specific guidance see CONSORT for harms[iii]) | | Results (Pickering, Amy J., et al. (2019))<br>Not updated for this study, as focused on secondary sample analysis (Montealegre et al.) |

| | | | | |
|---|---|---|---|---|
| **Discussion** | | | | |
| **Limitations** | 20 | Trial limitations, addressing sources of potential bias, imprecision, and, if relevant, multiplicity of analyses | | Discussion (Montealegre, Maria C., et al. (2021))<br>Discussion (Pickering, Amy J., et al. (2019)) |
| **Generalisability** | 21 | Generalisability (external validity, applicability) of the trial findings | Generalisability to clusters and/or individual participants (as relevant) | Discussion (Montealegre, Maria C., et al. (2021))<br>Discussion (Pickering, Amy J., et al. (2019)) |
| **Interpretation** | 22 | Interpretation consistent with results, balancing benefits and harms, and considering other relevant evidence | | Discussion (Montealegre, Maria C., et al. (2021))<br>Discussion (Pickering, Amy J., et al. (2019)) |
| **Other information** | | | | |
| **Registration** | 23 | Registration number and name of trial registry | | Methods (Pickering, Amy J., et al. (2019))<br>Not updated for this study (Montealegre et al.) |
| **Protocol** | 24 | Where the full trial protocol can be accessed, if available | | Published on Open Science Framework, link in Methods (Pickering, Amy J., et al. (2019))<br>Not updated for this study (Montealegre et al.) |
| **Funding** | 25 | Sources of funding and other support (such as supply of drugs), role of funders | | Summary, Funding (Montealegre, Maria C., et al. (2021))<br>Abstract, Role of Funder section (Pickering, Amy J., et al. (2019)) |

[1] Relevant to Conference Abstracts
